# Age at menopause and the risk of stroke: Observational and Mendelian Randomization analysis in 204,244 postmenopausal women

**DOI:** 10.1101/2023.02.08.23285676

**Authors:** Lena Tschiderer, Sanne AE Peters, Yvonne T van der Schouw, Anniek C van Westing, Tammy YN Tong, Peter Willeit, Lisa Seekircher, Conchi Moreno-Iribas, José María Huerta, Marta Crous-Bou, Martin Söderholm, Matthias B Schulze, Cecilia Johansson, Sara Själander, Alicia K Heath, Alessandra Macciotta, Christina C Dahm, Daniel B Ibsen, Valeria Pala, Lene Mellemkjær, Stephen Burgess, Angela Wood, Rudolf Kaaks, Verena Katzke, Pilar Amiano, Miguel Rodriguez-Barranco, Gunnar Engström, Elisabete Weiderpass, Anne Tjønneland, Jytte Halkjær, Salvatore Panico, John Danesh, Adam Butterworth, N Charlotte Onland-Moret

## Abstract

**Background:** Observational studies have shown that women with an early menopause are at higher risk of stroke compared to women with a later menopause. However, associations with stroke subtypes are inconsistent and the causality is unclear. Therefore, we conducted a large-scale analysis to investigate the observational association between age at menopause and different types of stroke accompanied by a Mendelian Randomization analysis to evaluate causality.

**Methods:** We analyzed data of the UK Biobank and EPIC-CVD study. Postmenopausal women without a history of stroke at baseline were eligible for inclusion. The study endpoints were total stroke and stroke subtypes (i.e., ischemic stroke, hemorrhagic stroke, intracerebral hemorrhage, and subarachnoid hemorrhage). We investigated the observational association between age at menopause and risk of stroke using Cox-regression analysis in each study separately before combining effect sizes using random-effects meta-analysis. Cox-regression analyses were progressively adjusted for (1) age, (2) smoking status, body mass index, glycated hemoglobin, total cholesterol, and hypertension, and (3) ever use of hormone replacement therapy and age at menarche. We used two-sample Mendelian Randomization analysis to study whether there is a causal relationship between genetically proxied age at menopause and risk of stroke.

**Results:** A total of 204,244 women were included (7,883 from EPIC-CVD [5,292 from the sub-cohort]; 196,361 from the UK Biobank). Pooled mean baseline age was 58.9 years (SD 5.8) and pooled mean age at menopause was 47.8 years (SD 6.2). Natural menopause occurred in 77.6% of all women. Over a median follow-up of 12.6 years (IQR 11.8, 13.3), 6,770 women experienced a stroke. In multivariable adjusted observational analyses, the pooled hazard ratios per five years younger age at menopause were 1.09 (95% CI: 1.07, 1.12) for stroke, 1.09 (1.06, 1.13) for ischemic stroke, 1.10 (1.04, 1.16) for hemorrhagic stroke, 1.14 (1.08, 1.20) for intracerebral hemorrhage, and 1.00 (0.84, 1.20) for subarachnoid hemorrhage. The Mendelian Randomization analysis found no evidence for a causal relationship between genetically proxied age at menopause and risk of any type of stroke.

**Conclusions:** Earlier age at menopause is associated with, but not causally related to the risk of stroke.

**Clinical Perspective:** **What is new?**

- This analysis involves over 200,000 postmenopausal women and more than 6,000 incident stroke cases and investigates the observational association between age at menopause and various subtypes of stroke. Furthermore, a Mendelian Randomization analysis was conducted to study whether associations are causal or not.
- Earlier age at menopause was statistically significantly associated with a higher risk of stroke and its subtypes ischemic stroke, hemorrhagic stroke, and intracerebral hemorrhage. We found no statistically significant relationship between earlier or later age at menopause and risk of subarachnoid hemorrhage.
- The Mendelian Randomization analysis suggested no causal effect of genetically proxied age at menopause and risk of any type of stroke.

**What are the clinical implications?**

- Women with earlier age at menopause are at higher risk of stroke. The underlying reasons need to be further investigated.
- Our analysis suggested that earlier menopause *per se* does not cause stroke. For prevention and adequate treatment of stroke in women, a better understanding of the specific role of menopause and the mechanistic background that leads to higher risk of stroke is needed.

## Introduction

Stroke is the second leading cause of death worldwide and was responsible for over six million deaths in 2019.^1^ At a global level, the proportion of deaths caused by stroke is higher for women (12.5% in 2019) than for men (10.9% in 2019).^2^ Women and men are also prone to different types of stroke. In a large-scale study in over nine million individuals, men had a higher risk of developing ischemic stroke, transient ischemic attack, and intracerebral hemorrhage, while women were at higher risk of subarachnoid hemorrhage.^3^ Women and men share several risk factors for stroke but the strengths of associations can differ between sexes.^4^ In addition, the relationship of various female-specific factors with cardiovascular risk has recently received increasing attention.^5^

The transition to menopause is predominantly defined by hormonal changes, and is accompanied by multi-faceted symptoms, such as sleep disturbances and vasomotor dysfunction.^6^ Recent data suggest a relationship between earlier menopause and the risk of developing cardiovascular disease.^7,8^ A large-scale individual participant data meta-analysis of the International Collaboration for a Life Course Approach to Reproductive Health and Chronic Disease Events (InterLACE) consortium found a higher risk of developing stroke in women with earlier age at natural menopause.^9^ The hazard ratio (HR) for stroke was 1.72 (95% confidence interval [CI]: 1.43, 2.07) for women who experienced menopause before the age of 40 years compared to women who experience menopause at age 50 or 51 years.^9^ The majority of previous studies on age at menopause and risk of stroke focused on a combined stroke endpoint or analyzed broader categories of ischemic and hemorrhagic stroke rather than specific stroke subtypes.

Since associations between age at menopause and risk of stroke have been based on data from observational studies, which are prone to confounding, the causality of the associations is unclear. Importantly, although earlier menopause is also associated with a higher risk of coronary heart disease,^9^ we showed in a recent Mendelian Randomization analysis that this association is unlikely to be causal.^10^ Furthermore, a Mendelian Randomization analysis based on publicly available results reported no significant causal relationship between genetically proxied age at natural menopause and risk of coronary artery disease or stroke.^11^ Similarly, a recently published study demonstrated no causal association between age at natural menopause and risk of ischemic stroke, although a small number of genetic variants was used.^12^ Whether this is also true for other types of stroke is still unclear.

We conducted a large-scale analysis including 204,244 postmenopausal women from the UK Biobank (UKB) and European Prospective Investigation into Cancer and Nutrition-Cardiovascular Diseases (EPIC-CVD) study to quantify the observational association between age at menopause and different types of stroke, and to investigate whether these associations were likely to be causal by applying a Mendelian Randomization analysis.

## Methods

### Study participants

In the present analysis, we included data from the UKB and EPIC-CVD study. Further details about these studies have been published previously.^13–16^ In brief, the UKB is a large-scale prospective study in the UK in which over 500,000 individuals aged 40 to 69 years were recruited between 2006 and 2010.^13,14^ The UKB was approved by the North West Multi-Centre Research Ethics Committee and all participants provided written informed consent. EPIC-CVD is a case-cohort study nested in the prospective cohort study EPIC, which recruited more than 500,000 individuals between 35 and 70 years of age in 1992-2000 from 23 centers throughout Europe.^15–17^ For the case-cohort study EPIC-CVD, a random sub-cohort was selected from the EPIC study for which a variety of biomarkers were obtained. In addition, EPIC-CVD includes all incident coronary heart disease and stroke events that occurred outside the sub-cohort. The EPIC study complies with the Declaration of Helsinki, and all participants gave written informed consent before participating. The study was approved by the local ethics committees of the participating centers and the Institutional Review Board of the International Agency for Research on Cancer (IARC, Lyon).

For the current analysis, postmenopausal women free of history of stroke at study baseline were eligible for inclusion. Furthermore, for EPIC-CVD, we excluded women with incident coronary heart disease and without incident stroke outside the sub-cohort due to the case-cohort design of study. **Figure 1** provides a flow chart on the selection of participants contributing to the current analysis. Of the 35,455 EPIC-CVD participants, we excluded 16,788 men, 632 women from French centers because follow-up for stroke was unavailable, 87 from Norway because important covariates were not measured, and 1,034 from Greece due to administrative constraints. Furthermore, we excluded 4,283 women with incident coronary heart disease outside the EPIC-CVD sub-cohort, 29 women with a history of stroke at baseline, and 4,719 women who were not postmenopausal, leaving 7,883 EPIC-CVD participants contributing to the current analysis. Of these 7,883 postmenopausal women, 5,292 belonged to the sub-cohort and 2,591 were stroke cases outside the sub-cohort (with 147 further stroke cases also belonging to the sub-cohort). Of the 502,412 individuals from UKB, 229,086 were excluded because they were male, 3,732 because they had a history of stroke at baseline, and 73,233 because they were not postmenopausal at baseline leaving 196,361 UKB participants contributing to our analysis. Consequently, we included a total of 204,244 postmenopausal women from both studies in the observational analysis.

**Figure 1.**
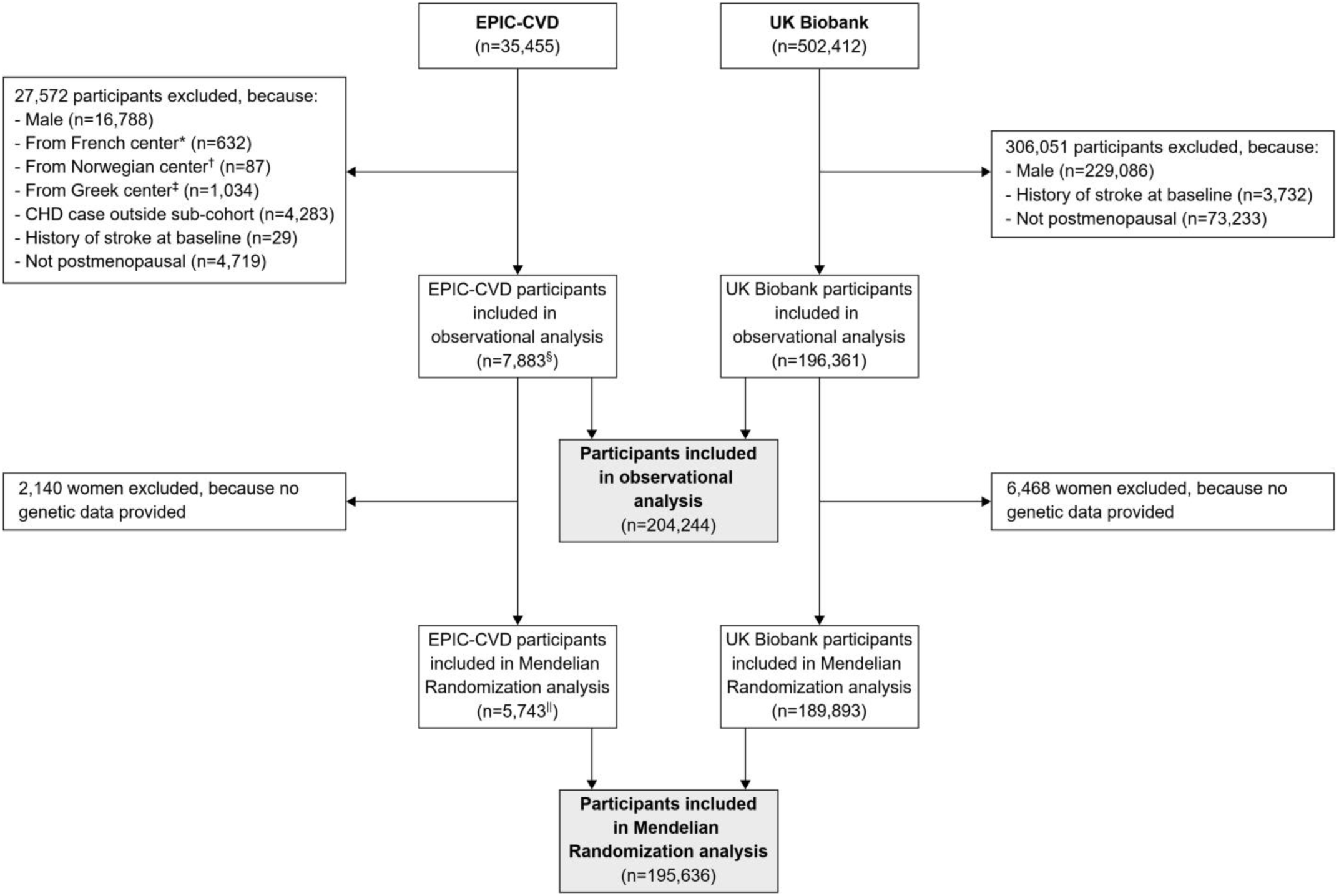
Participant flow chart. *Excluded because follow-up for stroke was unavailable in French centers. ^†^Excluded because important covariates were not measured in Norwegian centers. ^‡^Excluded because of administrative constraints. ^§^Of these, 5,292 belonged to the sub-cohort and 2,591 were stroke cases outside the sub-cohort. ^||^Of these, 4,127 belonged to the sub-cohort and 1,616 were stroke cases outside the sub-cohort.

### Definition of menopause, age at menopause, and type of menopause

Women were defined as being postmenopausal if they fulfilled at least one of the following criteria: (1) experienced natural menopause (defined as stopping of periods in UKB and as reporting no menses for one year or longer due to natural menopause in EPIC-CVD), (2) had had a unilateral or bilateral ovariectomy in EPIC-CVD or bilateral ovariectomy in UKB, or (3) had had a hysterectomy. Moreover, where no information on menopausal status was provided, we defined women aged >54 years as postmenopausal, as suggested previously.^10^ Type of menopause was defined as surgical if a history of ovariectomy or hysterectomy had been reported and as natural, otherwise. Age at menopause was defined as age of a woman’s last menstruation or, in case of a surgical menopause, the age at ovariectomy or hysterectomy.

### Outcome definition

We analyzed a combined stroke endpoint including fatal and non-fatal ischemic and hemorrhagic stroke with ICD-10 codes I60, I61, I63, and I64. Furthermore, we analyzed the individual stroke endpoints ischemic stroke (I63, I64), hemorrhagic stroke (I60, I61), intracerebral hemorrhage (I61), and subarachnoid hemorrhage (I60). For UKB participants, September 30, 2021, was used as end of follow-up for stroke. In EPIC-CVD, end of follow-up for stroke varied between the centers ranging from 2003 to 2010. Time to event was defined as time to stroke, death, or end of follow-up, whichever occurred first. For the analysis of stroke sub-types, we censored individual stroke events against each other and defined time to stroke as time to the first individual stroke endpoint. For instance, if an individual experienced both ischemic and hemorrhagic stroke, we only analyzed the first stroke event that occurred during follow-up. In case two types of stroke occurred on the same day, we gave preference to ischemic stroke over intracerebral hemorrhage over other types of stroke.

Details about the assessment and definition of additional variables used in our analyses are described in the **Supplementary Methods**.

### Genetic data

We obtained individual-level imputed data on genetic variants from both EPIC-CVD and UKB. Genotyping in EPIC-CVD was performed using the Human Core Exome array, Illumina 660 Quad array, and Omni Exome Express array.^18^ In the UKB, participants were genotyped with the Affymetrix UK BiLEVE Axiom array and the Affymetrix UKB Axiom Array.^14,19^ Genotype imputation was performed using the Haplotype Reference Consortium for EPIC-CVD^18^ and the Haplotype Reference Consortium as well as the UK10K haplotype reference panel for the UKB^20^.

For the Mendelian Randomization analysis, to quantify genetically proxied age at menopause, we used genetic variants reported to be associated with age at menopause by a large-scale genome-wide association study (GWAS) for our instrumental variable.^11^ A detailed selection process of the single nucleotide polymorphisms (SNPs) is described in the **Supplementary Methods**. Of the 290 SNPs identified by the GWAS, we excluded 124 SNPs because they were unavailable (n=63), palindromic (n=16), or rare with minor allele frequencies <0.1 (n=45), and used the remaining 166 SNPs to determine genetically proxied age at menopause in our main Mendelian Randomization analysis (see **eFigure 3** and **eTable 1**).

As shown in **Figure 1**, from the 204,244 postmenopausal women included in our observational analysis, we excluded 2,140 women from EPIC-CVD and 6,468 women from the UKB because data on the SNPs included in our analysis were unavailable, leaving 195,636 women included in our Mendelian Randomization analysis.

### Statistical analysis

#### Descriptive statistics

Summary statistics of continuous variables are provided as means and standard deviations (SDs) if normally distributed or as medians and interquartile ranges (IQRs), otherwise. Categorical variables are summarized as numbers and percentages. Pooled means across both cohorts were obtained from random-effects meta-analysis. To enhance comparability, we provide descriptive statistics including EPIC-CVD participants from the sub-cohort only.

#### Observational analyses

For the observational analysis, we estimated the association between age at menopause and incidence of stroke using Cox-regression analysis. For UKB, we implemented a Cox proportional hazards model. For EPIC-CVD, we additionally took into account the case-cohort design of the study by implementing Prentice-weighted Cox-regression analysis with robust standard errors.^21^ For both studies, we used age at baseline as the underlying time scale. We investigated the relationship between age at menopause and risk of stroke implementing age at menopause as a continuous variable and also by categorizing it as <40, 40 to <45, 45 to <50, 50 to <55, and ≥55 years, using 50 to <55 years at menopause as the reference category. When analyzing age at menopause as a categorical variable, we also obtained P-values for linear trends by treating the categorical variable as a continuous variable in our model. Furthermore, we present 95% CIs for each category using quasi variances to enhance the comparison between individual categories.^22^ When analyzing age at menopause as a continuous variable, we reported HRs per five years younger age at menopause. For graphical demonstration purposes and to assess non-linearity, we also analyzed age at menopause using restricted cubic splines with three knots at 45, 50, and 55 years of age at menopause using an age of 50 years at menopause as reference. We progressively adjusted our analysis for (1) age at baseline (Model 1), (2) smoking status (never, ex, current), body mass index (kg/m^2^), glycated hemoglobin (HbA1c, %), total cholesterol (mmol/L), and hypertension (yes, no) at baseline (Model 2), and (3) ever use of hormone replacement therapy (HRT) at baseline (yes, no) and age at menarche (years) (Model 3). For EPIC-CVD, we additionally stratified all models by country. Finally, we combined study-specific results using random-effects meta-analysis when including age at menopause as continuous variable, and multivariate random-effects meta-analysis^23^ when analyzing it as categorical variable. We also used multivariate random-effects meta-analysis^23^ to combine beta coefficients of the restricted cubic splines model. We imputed missing values using multiple imputation by chained equations with 14 datasets and 30 iterations (see **Supplementary Methods** for more details).

We estimated HRs for risk of stroke per five years younger age at menopause across the following subgroups: use of HRT (ever vs never), type of menopause (surgical vs natural), smoking status (current, ex vs never), and age at baseline. We included age at menopause, the subgroup variable of interest, and a formal interaction term between age at menopause and the subgroup variable of interest into our model. In addition, we adjusted the models for the baseline variables age, smoking status, body mass index, HbA1c, total cholesterol, hypertension, ever use of HRT, and age at menarche, if appropriate. For illustrative purposes we categorized age at baseline into <60 vs ≥60 years. The P-value for interaction for age at baseline was obtained from a model in which age at baseline was included as a continuous variable. For EPIC-CVD, we additionally stratified the model by country. We obtained effect estimates in each study and combined them using multivariate random-effects meta-analysis.^23^

#### Mendelian Randomization analysis

For the effect of the genetic variants on age at menopause, we used the beta coefficients and standard errors reported by the GWAS published by Ruth et al.^11^ without the UKB data in order to avoid sample overlap. We transformed beta coefficients and standard errors to reflect genetic effects per five years age at menopause to enhance comparability to our observational analysis. Then, we investigated the strength of our genetic instrument based on the F-statistic. For both EPIC-CVD and UKB, the F-statistic was calculated from linear regression including all genetic variants as independent variables and age at menopause as the dependent variable using the first imputed dataset and only including individuals from the sub-cohort for EPIC-CVD. Next, we estimated the effect of genetically proxied age at menopause and risk of different types of stroke using Cox-regression analysis with age as the underlying time scale. For UKB, we adjusted our model for age at baseline and the first 16 genetic principal components as suggested by Privé et al.^24^ For EPIC-CVD, we adjusted our model for age at baseline, the first ten genetic principal components (as 16 were not available), and genotype array, stratified by country, and implemented Prentice-weighted Cox-regression.^21^ We obtained effect estimates and standard errors for the association of each SNP with risk of different types of stroke. Finally, we conducted the Mendelian Randomization analysis based on (1) the effect estimates of the association between the SNPs and risk of different types of stroke we obtained from the UKB and EPIC-CVD and (2) the GWAS summary effect estimates on the relationship between the SNPs and age at menopause using the R-package *MendelianRandomization*.^25^ We applied standard inverse-variance weighted (IVW) regression to obtain the effect estimates for the association of genetically proxied age at menopause with risk of different types of stroke. We combined the effect sizes across studies using fixed-effect meta-analysis.

We conducted several sensitivity analyses. First, we applied different Mendelian Randomization analysis methods including simple median regression, weighted median regression, and MR-Egger regression. Second, we conducted a leave-one-out analysis omitting each SNP in turn to identify whether one of the SNPs particularly drives the result. Third, to study the distribution of stroke-related and female-specific factors across genetically proxied age at menopause, we obtained a polygenic risk score by calculating a weighted sum of the SNPs used in our analysis, weighting each SNP with the corresponding summary effect size obtained from the GWAS. We divided the polygenic risk score into study-combined fifths (fifths were comparable between the two studies) and compared the variables smoking status, body mass index, HbA1c, total cholesterol, hypertension, ever use of HRT, and age at menarche, across the five categories. Fourth, we further investigated pleiotropy by implementing MR-PRESSO.^26^ Fifth, to consider the effect of rare genetic variants, we excluded SNPs with minor allele frequencies <0.01 (n=8), rather than those with minor allele frequencies <0.1 as we did in our primary analysis. This sensitivity analysis included 203 SNPs, which are listed in **eTable 1**. Sixth, we additionally adjusted our Mendelian Randomization analysis for phenotypes associated with cardiovascular risk, i.e., smoking status, body mass index, HbA1c, total cholesterol, hypertension, ever use of HRT, and age at menarche.

Statistical analyses were carried out using R 4.0.5 (The R Foundation, Vienna, Austria). All statistical tests were two-sided and P-values ≤0.05 were deemed as statistically significant.

## Results

### Study population

Baseline characteristics of the study participants are demonstrated in **Table 1**, separately for the EPIC-CVD sub-cohort and UKB, and pooled across both cohorts. Overall, mean age was 58.9 years (SD 5.8) at baseline and 47.8 years (SD 6.2) at menopause. Surgical menopause was reported by 22.4% of the participants. Mean age at surgical and natural menopause was 41.9 (SD 7.0) and 49.8 (SD 4.6), respectively. Moreover, 51.3% of the women had a history of hypertension, 33.3% of the women smoked previously, and 8.7% smoked currently. Half of the women reported to have used HRT during their lifetime.

**Table 1.** Baseline characteristics.

| Characteristic | Overall<br>(n=201,653*) |  | EPIC-CVD<br>(n=5,292*) |  | UK Biobank<br>(n=196,361) |  |
| --- | --- | --- | --- | --- | --- | --- |
|  | Total<br>No. | Mean ± SD,<br>median [IQR],<br>No. (%†) | Total<br>No. | Mean ± SD,<br>median<br>[IQR], No.<br>(%†) | Total<br>No. | Mean ± SD,<br>median [IQR],<br>No. (%†) |
| **Stroke risk factors** |  |  |  |  |  |  |
| Age, years | 201,653 | 58.9 ± 5.8 | 5,292 | 58.0 ± 6.4 | 196,361 | 59.9 ± 5.7 |
| Systolic BP, mmHg | 189,007 | 137.1 ± 19.2 | 4,296 | 135.7 ± 20.0 | 184,711 | 138.4 ± 19.2 |
| Diastolic BP, mmHg | 189,009 | 81.6 ± 9.9 | 4,296 | 82.0 ± 10.4 | 184,713 | 81.2 ± 9.9 |
| Hypertension | 200,858 | 103,069 (51.3%) | 5,245 | 2,362 (45.0%) | 195,613 | 100,707 (51.5%) |
| Body mass index, kg/m<sup>2</sup> | 200,628 | 26.8 ± 5.1 | 5,258 | 26.4 ± 4.6 | 195,370 | 27.3 ± 5.1 |
| Education | 197,661 |  | 5,182 |  | 192,479 |  |
| Low |  | 43,350 (21.9%) |  | 2,026 (39.1%) |  | 41,324 (21.5%) |
| Medium |  | 48,764 (24.7%) |  | 615 (11.9%) |  | 48,149 (25.0%) |
| High |  | 104,994 (53.1%) |  | 1,988 (38.4%) |  | 103,006 (53.5%) |
| Smoking status | 200,560 |  | 5,254 |  | 195,306 |  |
| Never |  | 116,329 (58.0%) |  | 3,071 (58.5%) |  | 113,258 (58.0%) |
| Ex |  | 66,778 (33.3%) |  | 1,092 (20.8%) |  | 65,686 (33.6%) |
| Current |  | 17,453 (8.7%) |  | 1,091 (20.8%) |  | 16,362 (8.4%) |
| Total cholesterol, mmol/L | 188,029 | 6.2 ± 1.1 | 5,049 | 6.3 ± 1.2 | 182,980 | 6.0 ± 1.1 |
| HDL cholesterol, mmol/L | 171,617 | 1.6 ± 0.4 | 5,049 | 1.6 ± 0.4 | 166,568 | 1.6 ± 0.4 |
| Triglycerides, mmol/L | 187,916 | 1.4 [1.0, 2.0] | 5,046 | 1.2 [0.8, 1.6] | 182,870 | 1.4 [1.0, 2.0] |
| HbA1c, % | 186,905 | 5.6 ± 0.5 | 5,194 | 5.6 ± 0.7 | 181,711 | 5.5 ± 0.5 |
| **Female-specific factors** |  |  |  |  |  |  |
| Age at menopause, years | 189,095 | 47.8 ± 6.2 | 4,754 | 47.3 ± 6.4 | 184,341 | 48.4 ± 6.2 |
| Surgical menopause | 201,653 | 45,193 (22.4%) | 5,292 | 1,235 (23.3%) | 196,361 | 43,958 (22.4%) |
| Ever use of HRT | 200,206 | 100,203 (50.0%) | 4,722 | 1,733 (36.7%) | 195,484 | 98,470 (50.4%) |
| Ever use of OCP | 200,736 | 155,303 (77.4%) | 5,206 | 2,329 (44.7%) | 195,530 | 152,974 (78.2%) |
| Age at menarche, years | 195,496 | 13.2 ± 1.6 | 5,156 | 13.4 ± 1.6 | 190,340 | 12.9 ± 1.6 |

\*Only participants of the EPIC-CVD sub-cohort were included in this table. †The denominator for all percentages was the number of postmenopausal women without missing values in the respective variable. Abbreviations: BP, blood pressure; OCP, oral contraceptive pill; HbA1c, glycated hemoglobin; HRT, hormone replacement therapy; IQR, interquartile range; SD, standard deviation.

### Observational analysis

Over a median follow-up of 12.6 years (IQR 11.8, 13.3; 13.0 [10.8, 14.3] in EPIC-CVD and 12.6 [11.8, 13.3] in UKB), 6,770 women experienced a stroke. Of those, 5,155 were ischemic and 1,615 were hemorrhagic (976 intracerebral hemorrhages and 639 subarachnoid hemorrhages). Of the 6,770 strokes, 2,738 occurred in EPIC-CVD and 4,032 in UKB. Pooled results on the association of age at menopause with risk of stroke are provided in **Table 2** and **Figure 2**. Each five years younger at menopause was associated with a higher risk of total stroke (most adjusted HR 1.09 [95% CI: 1.07, 1.12]), ischemic stroke (1.09 [1.06, 1.13]), hemorrhagic stroke (1.10 [1.04, 1.16]), and intracerebral hemorrhage (1.14 [1.08, 1.20]). Age at menopause was not associated with the risk of subarachnoid hemorrhage (most adjusted HR 1.00 [0.84, 1.20] for each five years younger). Age at menopause was approximately log-linearly associated with the risk of stroke (P-value for trend <0.001), ischemic stroke (P-value for trend <0.001), hemorrhagic stroke (P-value for trend 0.022), and intracerebral hemorrhage (P-value for trend <0.001). For instance, compared to women who had experienced menopause between 50 and <55 years, multivariable adjusted HRs for stroke were 1.42 (1.28, 1.56), 1.23 (1.14, 1.33), 1.10 (1.02, 1.19), and 0.96 (0.84, 1.10) in women who had experienced menopause at ages <40, 40 to <45, 45 to <50, and ≥55 years, respectively. Separate results for EPIC-CVD and UKB are shown **eTable 2** and **eTable 3**, respectively. Results for all subtypes of stroke, except subarachnoid hemorrhage (higher risk for younger age at menopause in UKB but not in EPIC-CVD), were highly consistent between EPIC-CVD and UKB as demonstrated in **eFigure 1**.

**Figure 2.**
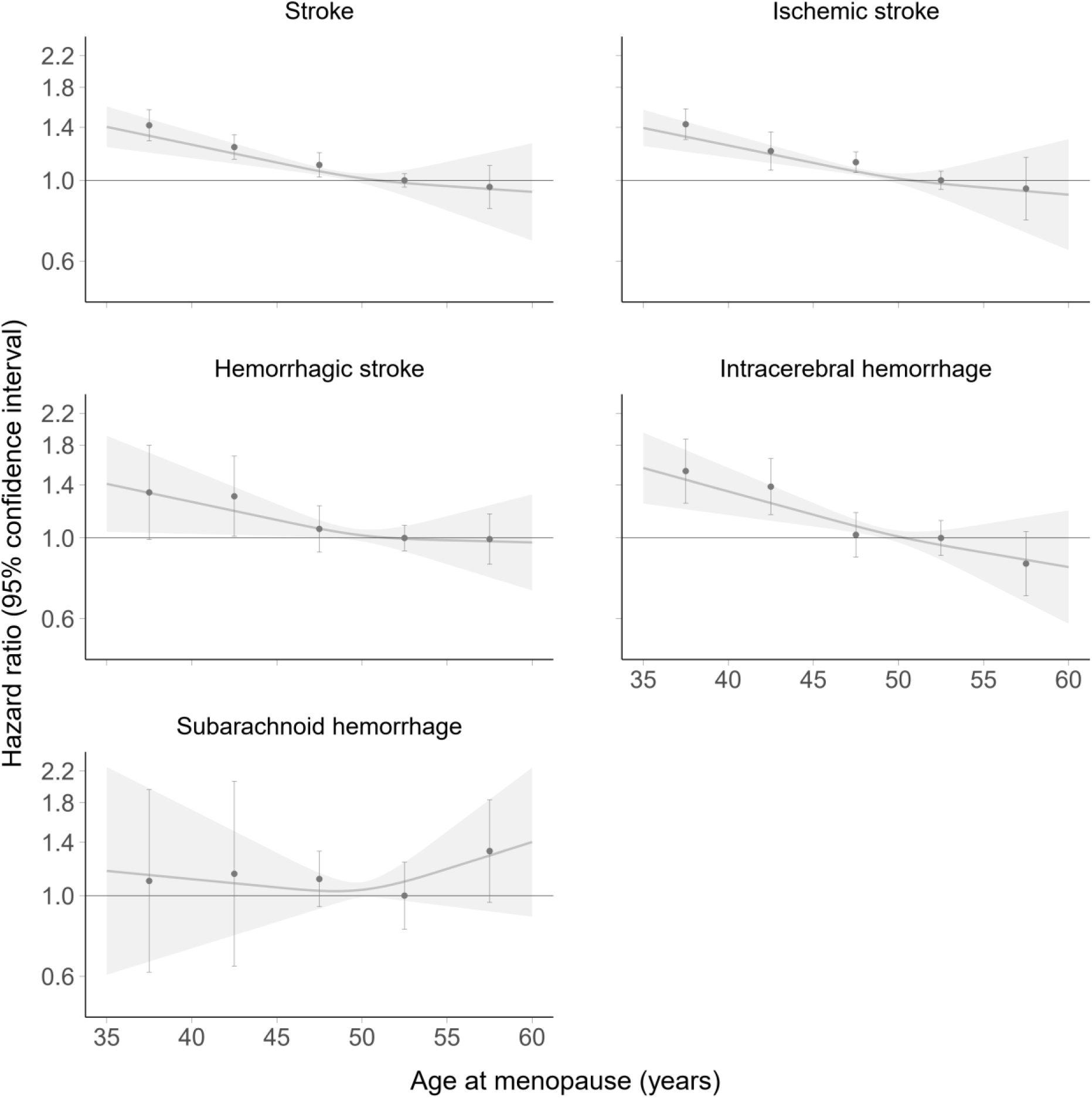
Association of age at menopause and risk of stroke. Hazard ratios are adjusted for age, smoking status, body mass index, glycated hemoglobin, total cholesterol, hypertension, ever use of hormone replacement therapy, and age at menarche. Dots indicate point estimates and whiskers 95% confidence intervals for the categories <40, 40 to <45, 45 to <50, 50 to <55, and ≥55 years of age at menopause. Age at menopause from 50 to <55 years was used as reference category. Restricted cubic splines are based on three knots at 45, 50, and 55 years of age at menopause using an age of 50 years at menopause as reference.

**Table 2.** Association between age at menopause and risk of stroke in the EPIC-CVD and UK Biobank studies (n=204,244).

| Outcome/Age at menopause (years) | No. of cases | Model 1 |  | Model 2 |  | Model 3 |  |
| --- | --- | --- | --- | --- | --- | --- | --- |
|  |  | HR (95% CI) | P<sub>trend</sub> | HR (95% CI) | P<sub>trend</sub> | HR (95% CI) | P<sub>trend</sub> |
| Stroke |  |  |  |  |  |  |  |
| <40 | 700 | 1.54 (1.36-1.75) |  | 1.41 (1.27-1.58) |  | 1.42 (1.28-1.56) |  |
| 40 to<45 | 891 | 1.31 (1.21-1.41) |  | 1.24 (1.15-1.33) |  | 1.23 (1.14-1.33) |  |
| 45 to <50 | 1,742 | 1.13 (1.05-1.23) | <0.001 | 1.10 (1.02-1.19) | <0.001 | 1.10 (1.02-1.19) | <0.001 |
| 50 to <55 | 2,623 | 1.00 (0.96-1.04) |  | 1.00 (0.96-1.04) |  | 1.00 (0.96-1.04) |  |
| ≥55 | 814 | 0.96 (0.83-1.10) |  | 0.95 (0.83-1.10) |  | 0.96 (0.84-1.10) |  |
| per 5 years younger | 6,770 | 1.12 (1.10-1.15) |  | 1.10 (1.07-1.12) |  | 1.09 (1.07-1.12) |  |
| Ischemic stroke |  |  |  |  |  |  |  |
| <40 | 534 | 1.59 (1.41-1.80) |  | 1.43 (1.28-1.58) |  | 1.43 (1.30-1.57) |  |
| 40 to<45 | 653 | 1.29 (1.15-1.45) |  | 1.21 (1.07-1.36) |  | 1.20 (1.07-1.36) |  |
| 45 to <50 | 1,350 | 1.16 (1.08-1.24) | <0.001 | 1.12 (1.05-1.20) | <0.001 | 1.12 (1.05-1.20) | <0.001 |
| 50 to <55 | 1,995 | 1.00 (0.95-1.05) |  | 1.00 (0.95-1.05) |  | 1.00 (0.94-1.06) |  |
| ≥55 | 623 | 0.95 (0.79-1.16) |  | 0.94 (0.77-1.15) |  | 0.95 (0.78-1.16) |  |
| per 5 years younger | 5,155 | 1.12 (1.10-1.15) |  | 1.09 (1.07-1.12) |  | 1.09 (1.06-1.13) |  |
| Hemorrhagic stroke |  |  |  |  |  |  |  |
| <40 | 166 | 1.37 (1.03-1.81) |  | 1.32 (0.97-1.79) |  | 1.33 (0.99-1.80) |  |
| 40 to<45 | 238 | 1.32 (1.03-1.69) |  | 1.29 (1.00-1.67) |  | 1.30 (1.01-1.68) |  |
| 45 to <50 | 392 | 1.08 (0.92-1.25) | 0.007 | 1.06 (0.91-1.22) | 0.036 | 1.06 (0.91-1.23) | 0.022 |
| 50 to <55 | 628 | 1.00 (0.93-1.08) |  | 1.00 (0.92-1.09) |  | 1.00 (0.92-1.08) |  |
| ≥55 | 191 | 0.97 (0.83-1.13) |  | 1.00 (0.85-1.17) |  | 0.99 (0.85-1.16) |  |
| per 5 years younger | 1,615 | 1.11 (1.06-1.17) |  | 1.09 (1.03-1.16) |  | 1.10 (1.04-1.16) |  |
| Intracerebral hemorrhage |  |  |  |  |  |  |  |
| <40 | 102 | 1.52 (1.25-1.86) |  | 1.50 (1.23-1.84) |  | 1.53 (1.25-1.87) |  |
| 40 to<45 | 145 | 1.38 (1.16-1.65) |  | 1.37 (1.15-1.64) |  | 1.38 (1.16-1.65) |  |
| 45 to <50 | 224 | 1.02 (0.88-1.17) | <0.001 | 1.02 (0.88-1.17) | <0.001 | 1.02 (0.89-1.17) | <0.001 |
| 50 to <55 | 394 | 1.00 (0.90-1.11) |  | 1.00 (0.90-1.12) |  | 1.00 (0.90-1.12) |  |
| ≥55 | 111 | 0.84 (0.69-1.03) |  | 0.85 (0.69-1.04) |  | 0.85 (0.69-1.04) |  |
| per 5 years younger | 976 | 1.15 (1.09-1.21) |  | 1.14 (1.08-1.20) |  | 1.14 (1.08-1.20) |  |
| Subarachnoid hemorrhage |  |  |  |  |  |  |  |
| <40 | 64 | 1.19 (0.71-2.00) |  | 1.08 (0.59-1.98) |  | 1.10 (0.62-1.96) |  |
| 40 to<45 | 93 | 1.20 (0.68-2.11) |  | 1.15 (0.63-2.08) |  | 1.15 (0.64-2.06) |  |
| 45 to <50 | 168 | 1.17 (0.94-1.45) | 0.615 | 1.11 (0.94-1.31) | 0.952 | 1.11 (0.93-1.32) | 0.923 |
| 50 to <55 | 234 | 1.00 (0.81-1.24) |  | 1.00 (0.81-1.24) |  | 1.00 (0.81-1.24) |  |
| ≥55 | 80 | 1.26 (0.96-1.65) |  | 1.33 (0.96-1.84) |  | 1.33 (0.96-1.83) |  |
| per 5 years younger | 639 | 1.03 (0.88-1.20) |  | 1.00 (0.82-1.21) |  | 1.00 (0.84-1.20) |  |

Model 1 is adjusted for age; Model 2 is additionally adjusted for smoking status, body mass index, HbA1c, total cholesterol, and hypertension; Model 3 is additionally adjusted for ever use of hormone replacement therapy and age at menarche. Confidence intervals for each age at menopause category are presented using quasi variances in order to enhance the comparison between individual categories. Age at menopause from 50 to <55 years was used as reference category. P<sub>trend</sub> indicates P-value for linear trend. Abbreviations: CI, confidence interval; HR, hazard ratio.

**eFigure 2** provides results of subgroup analyses for each individual stroke endpoint according to age at baseline, smoking status, use of HRT, and type of menopause. Age at baseline modified the association between age at menopause and total stroke (P-value for interaction 0.010) and ischemic stroke (P-value for interaction 0.003) with higher HRs for younger postmenopausal women. We did not find any statistically significant modification across all other subgroups (all P-values >0.05).

### Mendelian Randomization analysis

The F-statistics for the 166 SNPs included in our Mendelian Randomization analysis were 1.31 (R^2^: 5.2%) in EPIC-CVD and 23.2 (R^2^: 2.0%) in UKB.

Results of the Mendelian Randomization analysis are provided in **Figure 3** (see **eTable 4** for study-specific results). In contrast to the observational analysis, the IVW regression showed no evidence for a causal effect of age at menopause on stroke risk (HR per five years younger genetically proxied age at menopause 0.95 [95% CI: 0.82, 1.09; P-value=0.445]), nor for any of the individual stroke endpoints ischemic stroke, hemorrhagic stroke, intracerebral hemorrhage, or subarachnoid hemorrhage (all P-values>0.05).

**Figure 3.**
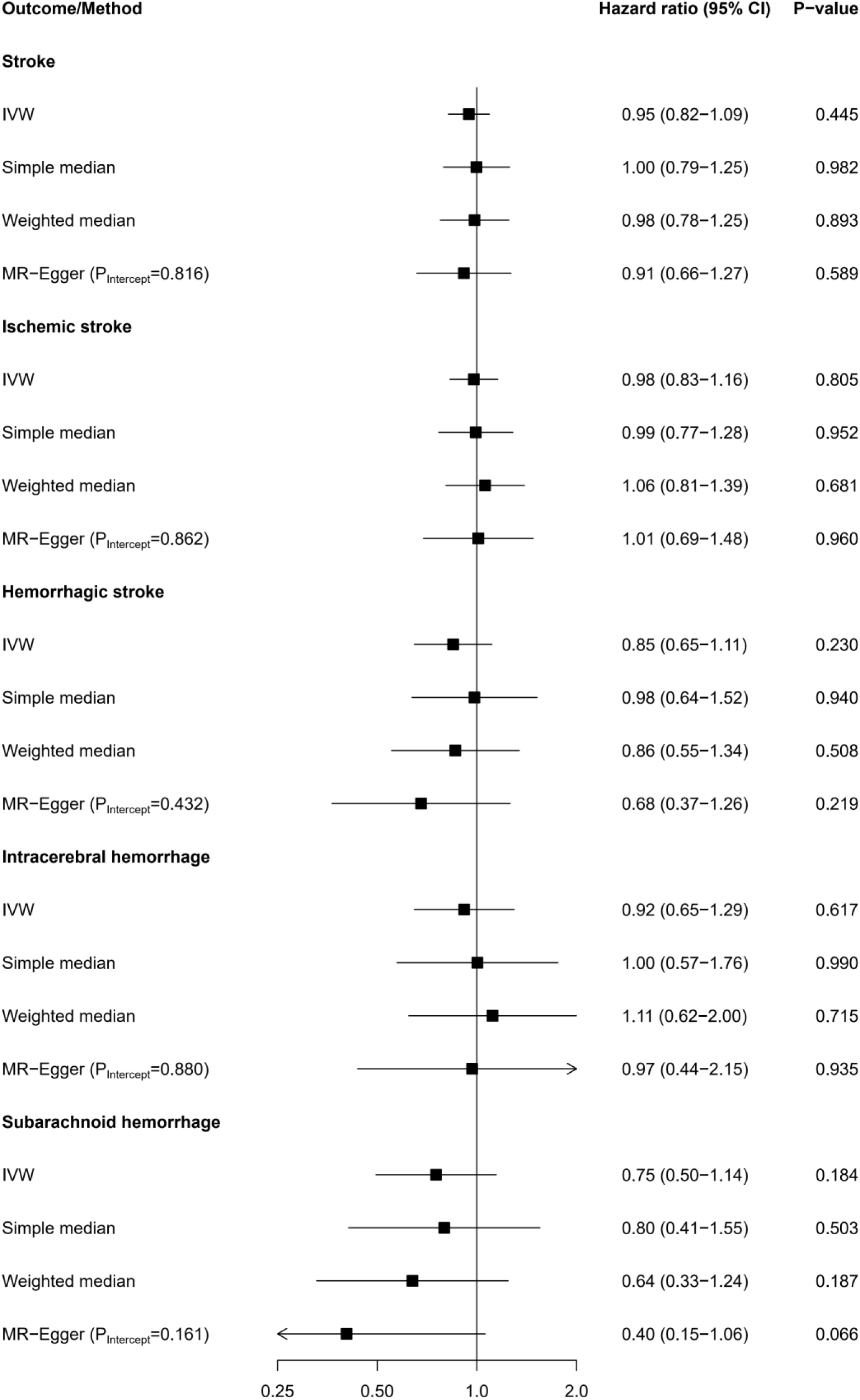
Mendelian Randomization analysis on genetically proxied age at menopause and risk of stroke. Hazard ratios are per five years younger genetically proxied age at menopause. Abbreviations: CI, confidence interval; IVW, inverse variance weighted.

### Sensitivity analyses

The results of the Mendelian Randomization analyses were similar when applying simple median, weighted median, and MR-Egger regression (see **Figure 3** and **eTable 4**). Furthermore, leave-one-out analyses showed robust associations when excluding each SNP in turn (data not shown). The MR-Egger method did not show any evidence for pleiotropy (P-values of intercept >0.05, see **Figure 3** and **eTable 4**). Furthermore, MR-PRESSO did not yield any SNPs that produced significant horizontal pleiotropy. After only excluding rare SNPs with minor allele frequencies <0.01 we again found no statistically significant relationship between genetically proxied age at menopause and risk of stroke as demonstrated in **eFigure 4**. Data on baseline patient characteristics across fifths of the polygenic risk score are provided in **eTable 5** (see distribution across age at menopause categories in **eTable 6**). There appeared to be an association between genetically proxied age at menopause and hypertension and ever use of HRT. However, when we adjusted our Mendelian Randomization analysis for all phenotypes, the association between genetically proxied age at menopause and risk of different types of stroke remained statistically nonsignificant as shown in **eFigure 5**.

## Discussion

In this large-scale analysis, we found an association between earlier age at menopause and higher risk of stroke, which was not likely to be causal according to Mendelian Randomization analysis.

### Strengths and limitations

Our study has several strengths. First, we included a large sample of postmenopausal women in our observational analyses and more than 6,000 incident stroke events providing excellent statistical power to detect associations and the potential influence of a variety of predefined, clinically relevant characteristics. Second, we investigated several subtypes of stroke including ischemic and hemorrhagic stroke and intracerebral and subarachnoid hemorrhage. Third, we meta-analyzed data from two large-scale studies and found highly consistent results for all stroke endpoints except subarachnoid hemorrhage, which may be explained by limited statistical power for this subtype. Fourth, we applied Mendelian Randomization analysis, a statistical method to study whether the results we found in our observational analyses were likely to be causal. Our analysis also has limitations. First, the EPIC-CVD and UKB studies had a different study design (case-cohort versus cohort study). To take this into account, we implemented Cox-regression with Prentice-weighting for EPIC-CVD. Second, the EPIC-CVD and UKB studies mainly include individuals of European ancestries (>95% for UKB), which limits the generalizability to other population groups. Of note, also the underlying GWAS focused on individuals of European ancestry. Consequently, more data on other populations are needed. Third, although we broadly harmonized the definition of menopause in EPIC-CVD and the UKB, it differed slightly between the studies as we did not have data on unilateral ovariectomy in the UKB. Fourth, menopausal status was missing for some women in the UKB and we estimated it based on age at baseline as suggested in previously published analyses on EPIC-CVD data.^27^ However, this only affected a small proportion of study participants (approximately 1% in UKB). Fifth, as age at menopause was self-reported, our analyses may also be affected by non-differential misclassification, which could have led to attenuation of the association towards the null. Sixth, although stroke events were ascertained comprehensively, there may remain some misclassification of events. However, it has been demonstrated that positive predictive values for stroke events defined based on hospital or death certificates are usually high.^28^ Furthermore, although we studied several types of stroke, stroke is a heterogeneous disease with many etiological subtypes and we were not able to study associations with more specific types of stroke. Seventh, Mendelian Randomization analysis relies on several assumptions. The first assumption is that the genetic variants are associated with the exposure variable, i.e., with age at menopause. We have used genetic variants published by a recent GWAS.^11^ After harmonization, we included 166 SNPs in our instrumental variable that had a sufficiently high F-statistic (i.e., >10 as suggested by Staiger and Stock^29^) for the UKB. In EPIC-CVD, the F-statistic was rather low indicating weak instrument bias, which is likely due to the lower sample size and large number of SNPs included (as the R^2^ statistic was even higher in EPIC-CVD than in the UKB). However, as we applied a two-sample Mendelian Randomization analysis, weak instrument bias acts towards the null and will therefore be conservative and we also found no significant effect of genetically proxied age at menopause on any type of stroke in the UKB. Moreover, the GWAS we used to define genetically proxied age at menopause focused on natural age at menopause. Therefore, our Mendelian Randomization study only tests for a causal association of age at natural menopause with stroke risk, while in our observational study we also included women with surgical menopause. However, our observational subgroup analyses showed no statistically significant difference in effect sizes between women with natural or surgical age at menopause. The second assumption of a Mendelian Randomization analysis relies on the principle that the genetic variants are not associated with confounding factors. We studied this assumption by comparing the polygenic risk score created by the SNPs included in our instrumental variable across several potential confounding factors and could not exclude an association between the polygenic risk score and hypertension and ever use of HRT. However, when we adjusted the Mendelian Randomization analysis for all phenotypes related to cardiovascular risk, the association remained nonsignificant. The third assumption of a Mendelian Randomization analysis is that the effect of the genetic variants on the outcome only goes through the exposure variable. To investigate this assumption, we (1) conducted a Mendelian Randomization analysis using MR-Egger, which yielded no significant intercepts, suggesting no evidence of directional pleiotropy, and (2) implemented MR-PRESSO, which resulted in no pleiotropic SNPs that should be excluded from the analysis.

### Comparison to previous observational studies

The results from our observational analysis on risk of total stroke are comparable to previous findings from individual studies including the Korean Heart Study,^30^ the China Kadoorie Biobank,^31^ and the Nurses’ Health Study^32^. The InterLACE consortium meta-analyzed data from over 300,000 women, whereby a large proportion of these data (61%) were from the UKB, leading to an overlap with our study sample.^9^ In analyses excluding data from the UKB, they also found an increased risk of developing stroke for women with younger age at menopause.^9^ Similarly, earlier age at menopause was associated with higher risk of ischemic stroke in the Korean Heart Study^30^ and Framingham Study.^33^ A study in textile workers from China did not confirm this association.^34^ However, in the InterLACE consortium, early menopause was related to higher risk of ischemic stroke, although these data included UKB as well.^9^ In our analysis, earlier menopause was also associated with a higher risk of hemorrhagic stroke. In contrast, two previous studies^30,34^ reported no significant association between age at menopause and risk of hemorrhagic stroke. This discrepancy is most likely due to limited statistical power, as within the InterLACE consortium, again including UKB, risk of hemorrhagic stroke was higher in women with earlier menopause.^9^ Finally, in our meta-analysis we found no significant relationship between age at menopause and risk of subarachnoid hemorrhage, unlike an investigation in the Nurses’ Health Study that reported a higher risk of aneurysmal subarachnoid hemorrhage for women with early menopause.^35^ Of note, our observational analysis in the UKB also found women with earlier age at menopause to be at higher risk of subarachnoid hemorrhage. However, after meta-analyzing data from the UKB with EPIC-CVD, the pooled hazard ratios were no longer statistically significant. This could probably be due to limited statistical power in EPIC-CVD as already mentioned above.

### Mendelian Randomization analysis

Our Mendelian Randomization analysis did not provide evidence that the associations found in observational analyses are likely to be causal. This finding is in line with a previous study on the relationship between reproductive aging and risk of coronary heart disease, which also found that genetically proxied reproductive aging had no causal effect on risk of coronary heart disease.^10^ Furthermore, a Mendelian Randomization analysis relying on publicly available GWAS results reported no significant causal relationship between genetically proxied age at natural menopause and risk of coronary artery disease or stroke.^11^ Similarly, a recently published Mendelian Randomization study, using fewer genetic variants, also reported that there is likely to be no causal link between age at natural menopause and risk of ischemic stroke.^12^ It is, therefore, likely that residual confounding is present in the observational analysis even though most observational studies controlled for potential confounding factors by adjusting for a set of stroke-related risk factors and female-specific factors. Identifying such confounding factors could help to better understand the development and progression of cardiovascular disease in women and might provide clues for previously unknown factors associated with menopause as well as stroke.

### Implications of the findings

Although our study did not find evidence of a potential causal relationship between age at menopause and risk of stroke, age at menopause may still be an important marker for cardiovascular disease in women as demonstrated in our observational analysis. Apparently, earlier age at menopause does not *per se* cause stroke but it captures an impact on the risk of developing stroke that may be determined by one or more confounding factors. This seems to hold for various types of stroke including both ischemic and hemorrhagic stroke. While there was a consistent association between earlier age at menopause and higher risk of intracerebral hemorrhage in both studies included in our meta-analysis, the relationship with higher risk of subarachnoid hemorrhage was only statistically significant in the UKB. As we were not able to account for the unknown factors driving the observational results in our analysis, further investigations are warranted. Several factors have been hypothesized to be responsible for the increased cardiovascular risk after menopause. One frequently discussed factor that has been speculated to have a cardioprotective effect in premenopausal women is estrogen.^36^ However, it has been shown that the association between estradiol and risk of myocardial infarction attenuated upon adjustment of age and other cardiovascular risk factors.^37^ Furthermore, when studying causal relations of age at menopause with different types of outcomes, associations appear to be specifically confounded when assessing cardiovascular risk as, for instance, genetically proxied age at menopause was causally related to risk of osteoporosis, fractures, and lung cancer.^12^ Understanding the mechanisms that lead to the observational relationship between earlier menopause and higher risk of stroke may help close an important knowledge gap that could enable us to better understand sex-differences in the development of stroke.

## Conclusion

Earlier age at menopause is associated with, but not causally related to, the risk of stroke.

## Data Availability

This research has been conducted using the UKB Resource (Application Number 29916). The UK Biobank resources are available upon reasonable request and can be accessed through applications on their website (https://www.ukbiobank.ac.uk/enable-your-research/apply-for-access). Information on data access for the EPIC study is provided at https://epic.iarc.fr/access/index.php. Summary-level GWAS have been published previously (doi: 10.1038/s41586-021-03779-7).

## Non-standard Abbreviations and Acronyms

CI: Confidence interval
EPIC-CVD: European Prospective Investigation into Cancer and Nutrition- Cardiovascular Diseases
GWAS: Genome-wide association study
InterLACE: International Collaboration for a Life Course Approach to Reproductive Health and Chronic Disease Events
IQR: Interquartile range
IVW: Inverse variance weighted
HbA1c: Glycated hemoglobin
HR: Hazard ratio
HRT: Hormone replacement therapy
SD: Standard deviation
SNP: Single nucleotide polymorphism
UKB: UK Biobank

## Acknowledgements

This research has been conducted using the UKB Resource (Application Number 29916). We are extremely grateful to all participants in UKB. This work was supported by core funding from the British Heart Foundation (RG/13/13/30194; RG/18/13/33946) and NIHR Cambridge Biomedical Research Centre (BRC-1215-20014) [*]. EPIC-CVD was funded by the European Research Council (268834), the European Commission Framework Programme 7 (HEALTH-F2-2012-279233) and Novartis. We thank all EPIC participants and staff for their contribution to the study, the laboratory teams at the Medical Research Council Epidemiology Unit for sample management and Cambridge Genomic Services for genotyping, Sarah Spackman for data management, and the team at the EPIC-CVD Coordinating Centre for study coordination and administration. *The views expressed are those of the author(s) and not necessarily those of the NIHR or the Department of Health and Social Care. The GWAS array was funded by the European Commission Framework Programme 7 (HEALTH-F2-2012-279233). Exomechip genotyping projects were funded by Pfizer and Merck. Where authors are identified as personnel of the International Agency for Research on Cancer / World Health Organization, the authors alone are responsible for the views expressed in this article and they do not necessarily represent the decisions, policy or views of the International Agency for Research on Cancer / World Health Organization. Furthermore, we thank Bas B L Penning de Vries for valuable advice on the multiple imputation analysis.

## Supplemental Material

Supplemental Methods

eTables 1-6

eFigures 1-5

## SUPPLEMENTAL MATERIAL

### Supplemental Methods

#### Assessment and definition of additional variables

In the UK Biobank, hypertension was defined as self-reported intake of antihypertensive medication or as having a systolic blood pressure >140 mmHg or a diastolic blood pressure >90 mmHg taking into account the average of two blood pressure measures taken a few moments apart. In EPIC-CVD, hypertension was defined as having a systolic blood pressure >140 mmHg, a diastolic blood pressure >90 mmHg, a self-reported history of hypertension, or as self-reported intake of antihypertensive medication. In both studies, body mass index was calculated by dividing weight in kg by height in m^2^. In EPIC-CVD, low socioeconomic status was defined according to education, whereby no or primary schooling indicated low, secondary schooling indicated medium, and vocational schooling or university indicated high socioeconomic status. In the UK Biobank, socioeconomic status was defined according to thirds of the Townsend index, whereby values ≤-3.2 indicated low, >-3.2 and ≤-0.803 indicated medium, and >-0.803 indicated high socioeconomic status. Smoking was divided into never, ex, and current tobacco smoking. Lipid levels, including total cholesterol, high-density lipoprotein cholesterol, and triglycerides, were measured in serum samples at baseline on a Roche auto-analyzer at Stichting Huisartsen Laboratorium (Etten-Leur, The Netherlands) in EPIC-CVD and using a Beckman Coulter AU5800 platform in UK Biobank. Erythrocyte glycated hemoglobin (HbA1c) was measured using the Tosoh-G8 HPLC analyzer (Tosoh Bioscience, Japan) in EPIC-CVD and with Bio-Rad Variant II Turbo analyzers in the UK Biobank. In the UK Biobank, HbA1c was transformed from mmol/mol to % in using the formula [HbA1c in %] = 2.15 + 0.0915*[HbA1c in mmol/mol].^38^ Physical activity was defined according to the Cambridge physical activity index in EPIC-CVD^39^ and according to the International Physical Activity Questionnaire (IPAQ) scoring protocol (short forms)^40^ in the UK Biobank. In addition, ever use of hormone replacement therapy and ever use of oral contraceptive pill was self-reported in both studies. Finally, age at menarche was defined as age at first menstrual period, which was self-reported in both studies.

#### Multiple imputation of missing values

We imputed missing values using multiple imputation by chained equations with 14 datasets and 30 iterations. We included the following variables into our imputation model: total cholesterol, high-density lipoprotein cholesterol, triglycerides, apolipoprotein A1, apolipoprotein B, HbA1c, ever use of oral contraceptive pill, age at menarche, age at menopause, body mass index, history of diabetes mellitus, hypertension, ever use of hormone replacement therapy, smoking status, age at baseline, menopausal status, physical activity, type of menopause, incident stroke indicator, incident ischemic stroke indicator, incident hemorrhagic stroke indicator, incident intracerebral hemorrhage indicator, incident subarachnoid hemorrhage indicator, the Nelson-Aalen estimator for stroke (weighted by the sampling fraction for EPIC-CVD)^41^ for both the UK Biobank and EPIC-CVD. Additionally, we used the variables systolic blood pressure, diastolic blood pressure, and Townsend index for the UK Biobank and the variables center, country, and education for EPIC-CVD. In case the Pearson correlation coefficient between two predictors was higher than 0.7 or lower than -0.7, we only used the predictor with the higher correlation coefficient with the imputed variable. Furthermore, we implemented passive multiple imputation for the variable representing categories of age at menopause. For the meta-analysis we used within-study multiple imputation as suggested by Burgess et al.^42^, i.e., we multiply imputed missing values in each study, combined the results by Rubin’s rule, and applied random-effects meta-analysis to combine the individual study results.

#### Selection of SNPs

The selection process of SNPs included in the Mendelian Randomization analysis is demonstrated in **eFigure 3**. A detailed list of SNPs included is provided in **eTable 1**. Of the 290 SNPs identified by the GWAS,^11^ we excluded (1) 63 SNPs because data were not available in EPIC-CVD; (2) 16 SNPs because they were palindromic and had an allele frequency between 0.3 and 0.7 obtained from either the summary data from the GWAS, within UK Biobank, or EPIC-CVD; and (3) 45 SNPs because they were rare with a minor allele frequency <0.1. Finally, we included 166 SNPs into our instrumental variable to genetically proxy age at menopause. Additionally, we checked whether the genetic variants were reported on the correct allele and harmonized the data accordingly.

### Supplemental Tables

**eTable 1.** Selection of SNPs.

| SNPs obtained from GWAS | SNPs in main analysis after exclusion, because |  |  | SNPs in sensitivity analysis after exclusion, because |
| --- | --- | --- | --- | --- |
|  | data not available in EPIC-CVD | palindromic and allele frequency between 0.3 and 0.7 | minor allele frequency <0.1 | minor allele frequency <0.01 |
| rs200448 | rs200448 | rs200448 | rs200448 | rs200448 |
| rs9438982 | rs9438982 | rs9438982 | rs9438982 | rs9438982 |
| rs12046563 | rs12046563 | rs12046563 | rs12046563 | rs12046563 |
| rs57259875 | rs57259875 | rs57259875 | rs57259875 | rs57259875 |
| rs112869704 | rs112869704 | rs112869704 | rs112869704 | rs112869704 |
| rs12133213 | — | — | — | — |
| rs11102802 | rs11102802 | rs11102802 | rs11102802 | rs11102802 |
| rs72708144 | rs72708144 | rs72708144 | — | rs72708144 |
| rs2736609 | rs2736609 | rs2736609 | rs2736609 | rs2736609 |
| rs41272475 | rs41272475 | rs41272475 | — | — |
| rs371125724 | — | — | — | — |
| rs11582336 | rs11582336 | rs11582336 | rs11582336 | rs11582336 |
| rs1044595 | rs1044595 | rs1044595 | rs1044595 | rs1044595 |
| rs7515939 | rs7515939 | — | — | — |
| rs4653680 | — | — | — | — |
| rs7414807 | rs7414807 | rs7414807 | rs7414807 | rs7414807 |
| rs1635506 | rs1635506 | rs1635506 | rs1635506 | rs1635506 |
| rs72755295 | rs72755295 | rs72755295 | — | rs72755295 |
| rs851797 | rs851797 | rs851797 | rs851797 | rs851797 |
| rs7539755 | rs7539755 | rs7539755 | rs7539755 | rs7539755 |
| rs7779 | rs7779 | rs7779 | — | rs7779 |
| rs112866529 | rs112866529 | rs112866529 | rs112866529 | rs112866529 |
| rs780088 | rs780088 | rs780088 | rs780088 | rs780088 |
| rs12053063 | rs12053063 | rs12053063 | rs12053063 | rs12053063 |
| rs10198789 | rs10198789 | rs10198789 | rs10198789 | rs10198789 |
| rs67600408 | rs67600408 | rs67600408 | — | rs67600408 |
| rs17425341 | rs17425341 | rs17425341 | rs17425341 | rs17425341 |
| rs76928871 | rs76928871 | rs76928871 | rs76928871 | rs76928871 |
| rs2293269 | rs2293269 | — | — | — |
| rs199638788 | — | — | — | — |
| rs62156756 | rs62156756 | rs62156756 | rs62156756 | rs62156756 |
| rs4852777 | rs4852777 | — | — | — |
| rs67893326 | rs67893326 | rs67893326 | rs67893326 | rs67893326 |
| rs59854799 | — | — | — | — |
| rs6727266 | rs6727266 | rs6727266 | rs6727266 | rs6727266 |
| rs72827480 | rs72827480 | rs72827480 | rs72827480 | rs72827480 |
| rs6430545 | rs6430545 | — | — | — |
| rs16830019 | rs16830019 | rs16830019 | rs16830019 | rs16830019 |
| rs4668354 | rs4668354 | — | — | — |
| rs201478184 | — | — | — | — |
| rs6719012 | rs6719012 | rs6719012 | rs6719012 | rs6719012 |
| rs4972504 | rs4972504 | rs4972504 | rs4972504 | rs4972504 |
| rs72934556 | rs72934556 | rs72934556 | rs72934556 | rs72934556 |
| rs540836853 | — | — | — | — |
| rs6736096 | rs6736096 | rs6736096 | rs6736096 | rs6736096 |
| rs7558434 | rs7558434 | rs7558434 | rs7558434 | rs7558434 |
| rs62193239 | rs62193239 | rs62193239 | rs62193239 | rs62193239 |
| rs606920 | rs606920 | rs606920 | rs606920 | rs606920 |
| rs12636454 | rs12636454 | rs12636454 | rs12636454 | rs12636454 |
| rs62244773 | rs62244773 | — | — | — |
| rs12487736 | rs12487736 | rs12487736 | rs12487736 | rs12487736 |
| rs1264191 | rs1264191 | rs1264191 | rs1264191 | rs1264191 |
| rs34675186 | — | — | — | — |
| rs9968117 | rs9968117 | rs9968117 | rs9968117 | rs9968117 |
| rs2885255 | rs2885255 | — | — | — |
| rs7610102 | — | — | — | — |
| rs4679244 | rs4679244 | rs4679244 | rs4679244 | rs4679244 |
| rs6793835 | rs6793835 | rs6793835 | rs6793835 | rs6793835 |
| rs1545597 | rs1545597 | rs1545597 | rs1545597 | rs1545597 |
| rs10154963 | rs10154963 | rs10154963 | rs10154963 | rs10154963 |
| rs344018 | rs344018 | rs344018 | rs344018 | rs344018 |
| rs72268838 | — | — | — | — |
| rs10804920 | rs10804920 | rs10804920 | rs10804920 | rs10804920 |
| rs979481644 | — | — | — | — |
| rs13070791 | rs13070791 | rs13070791 | rs13070791 | rs13070791 |
| rs3796624 | rs3796624 | rs3796624 | rs3796624 | rs3796624 |
| rs2052160 | rs2052160 | rs2052160 | rs2052160 | rs2052160 |
| rs190043078 | — | — | — | — |
| rs6824237 | rs6824237 | rs6824237 | rs6824237 | rs6824237 |
| rs2581458 | rs2581458 | rs2581458 | rs2581458 | rs2581458 |
| rs76540949 | — | — | — | — |
| rs12651246 | rs12651246 | rs12651246 | rs12651246 | rs12651246 |
| rs7698733 | — | — | — | — |
| rs145615497 | rs145615497 | rs145615497 | — | rs145615497 |
| rs6810489 | rs6810489 | rs6810489 | rs6810489 | rs6810489 |
| rs35713108 | — | — | — | — |
| rs112898082 | rs112898082 | rs112898082 | rs112898082 | rs112898082 |
| rs2139178 | rs2139178 | rs2139178 | rs2139178 | rs2139178 |
| rs9990489 | rs9990489 | rs9990489 | rs9990489 | rs9990489 |
| rs274701 | rs274701 | rs274701 | rs274701 | rs274701 |
| rs80167918 | rs80167918 | rs80167918 | — | rs80167918 |
| rs62356073 | rs62356073 | rs62356073 | rs62356073 | rs62356073 |
| rs17206591 | rs17206591 | rs17206591 | rs17206591 | rs17206591 |
| rs7728833 | rs7728833 | rs7728833 | rs7728833 | rs7728833 |
| rs10070308 | rs10070308 | rs10070308 | rs10070308 | rs10070308 |
| rs10477172 | rs10477172 | rs10477172 | rs10477172 | rs10477172 |
| rs888694 | rs888694 | rs888694 | — | rs888694 |
| rs11744866 | rs11744866 | rs11744866 | rs11744866 | rs11744866 |
| rs35813302 | rs35813302 | rs35813302 | rs35813302 | rs35813302 |
| rs2241584 | rs2241584 | rs2241584 | rs2241584 | rs2241584 |
| rs353478 | rs353478 | rs353478 | rs353478 | rs353478 |
| rs76623841 | rs76623841 | rs76623841 | — | rs76623841 |
| rs9348724 | rs9348724 | rs9348724 | rs9348724 | rs9348724 |
| rs72823382 | rs72823382 | rs72823382 | rs72823382 | rs72823382 |
| rs4716056 | rs4716056 | rs4716056 | rs4716056 | rs4716056 |
| rs9348752 | rs9348752 | rs9348752 | — | rs9348752 |
| rs6930435 | rs6930435 | rs6930435 | rs6930435 | rs6930435 |
| rs3130040 | rs3130040 | rs3130040 | — | rs3130040 |
| rs4143333 | rs4143333 | rs4143333 | — | rs4143333 |
| rs2844466 | rs2844466 | rs2844466 | rs2844466 | rs2844466 |
| rs9272211 | — | — | — | — |
| rs9272982 | — | — | — | — |
| rs7748483 | rs7748483 | rs7748483 | — | rs7748483 |
| rs9470134 | — | — | — | — |
| rs515650 | — | — | — | — |
| rs113967617 | rs113967617 | rs113967617 | rs113967617 | rs113967617 |
| rs2147220 | rs2147220 | rs2147220 | rs2147220 | rs2147220 |
| rs6569648 | rs6569648 | rs6569648 | rs6569648 | rs6569648 |
| rs140381358 | — | — | — | — |
| rs67149731 | — | — | — | — |
| rs61065130 | — | — | — | — |
| rs11767307 | rs11767307 | rs11767307 | rs11767307 | rs11767307 |
| rs1826838 | rs1826838 | rs1826838 | rs1826838 | rs1826838 |
| rs62445870 | rs62445870 | rs62445870 | — | rs62445870 |
| rs375846854 | — | — | — | — |
| rs10255049 | rs10255049 | rs10255049 | rs10255049 | rs10255049 |
| rs2023778 | rs2023778 | rs2023778 | rs2023778 | rs2023778 |
| rs2056726 | rs2056726 | rs2056726 | rs2056726 | rs2056726 |
| rs2392836 | rs2392836 | rs2392836 | rs2392836 | rs2392836 |
| rs4731541 | rs4731541 | — | — | — |
| rs112190116 | — | — | — | — |
| rs2013 | rs2013 | rs2013 | rs2013 | rs2013 |
| rs2061834 | rs2061834 | rs2061834 | rs2061834 | rs2061834 |
| rs3750243 | rs3750243 | rs3750243 | rs3750243 | rs3750243 |
| rs6473979 | rs6473979 | rs6473979 | — | rs6473979 |
| rs3735828 | — | — | — | — |
| rs73264416 | rs73264416 | rs73264416 | — | rs73264416 |
| rs35529153 | — | — | — | — |
| rs117431243 | — | — | — | — |
| rs1467044 | rs1467044 | rs1467044 | rs1467044 | rs1467044 |
| rs6470598 | rs6470598 | rs6470598 | rs6470598 | rs6470598 |
| rs1476164 | rs1476164 | rs1476164 | rs1476164 | rs1476164 |
| rs55873183 | rs55873183 | rs55873183 | — | rs55873183 |
| rs783562 | rs783562 | rs783562 | rs783562 | rs783562 |
| rs4879656 | rs4879656 | rs4879656 | rs4879656 | rs4879656 |
| rs10818873 | rs10818873 | rs10818873 | — | rs10818873 |
| rs138114862 | — | — | — | — |
| rs10795520 | rs10795520 | rs10795520 | rs10795520 | rs10795520 |
| rs74701710 | rs74701710 | rs74701710 | — | rs74701710 |
| rs10764106 | rs10764106 | rs10764106 | rs10764106 | rs10764106 |
| rs10997819 | rs10997819 | rs10997819 | rs10997819 | rs10997819 |
| rs10823203 | rs10823203 | rs10823203 | rs10823203 | rs10823203 |
| rs1889921 | rs1889921 | rs1889921 | rs1889921 | rs1889921 |
| rs7087644 | rs7087644 | rs7087644 | — | rs7087644 |
| rs74430332 | rs74430332 | rs74430332 | — | rs74430332 |
| rs10883451 | rs10883451 | rs10883451 | rs10883451 | rs10883451 |
| **rs7091889** | rs7091889 | rs7091889 | rs7091889 | rs7091889 |
| rs11245450 | rs11245450 | rs11245450 | rs11245450 | rs11245450 |
| rs728900 | rs728900 | — | — | — |
| rs6578283 | rs6578283 | rs6578283 | rs6578283 | rs6578283 |
| rs7928823 | — | — | — | — |
| rs34591978 | — | — | — | — |
| rs11031006 | rs11031006 | rs11031006 | rs11031006 | rs11031006 |
| rs10734411 | rs10734411 | rs10734411 | rs10734411 | rs10734411 |
| rs10769315 | rs10769315 | rs10769315 | rs10769315 | rs10769315 |
| rs7943302 | rs7943302 | rs7943302 | rs7943302 | rs7943302 |
| rs10899493 | rs10899493 | rs10899493 | rs10899493 | rs10899493 |
| rs11400191 | — | — | — | — |
| rs60378595 | — | — | — | — |
| rs148681796 | rs148681796 | rs148681796 | — | rs148681796 |
| rs10743271 | — | — | — | — |
| rs2376898 | rs2376898 | rs2376898 | rs2376898 | rs2376898 |
| rs77100210 | rs77100210 | rs77100210 | — | rs77100210 |
| rs61913600 | rs61913600 | rs61913600 | rs61913600 | rs61913600 |
| rs10743724 | rs10743724 | rs10743724 | rs10743724 | rs10743724 |
| rs10219645 | — | — | — | — |
| rs2277339 | rs2277339 | rs2277339 | rs2277339 | rs2277339 |
| rs28473132 | — | — | — | — |
| rs7308068 | rs7308068 | rs7308068 | rs7308068 | rs7308068 |
| rs75770066 | rs75770066 | rs75770066 | — | rs75770066 |
| rs61069255 | rs61069255 | rs61069255 | rs61069255 | rs61069255 |
| rs192638119 | — | — | — | — |
| rs5800506 | — | — | — | — |
| rs6490269 | rs6490269 | rs6490269 | rs6490269 | rs6490269 |
| rs2942371 | rs2942371 | rs2942371 | rs2942371 | rs2942371 |
| rs73413716 | rs73413716 | rs73413716 | — | rs73413716 |
| rs12825762 | rs12825762 | rs12825762 | rs12825762 | rs12825762 |
| rs35067339 | — | — | — | — |
| rs28416520 | rs28416520 | rs28416520 | rs28416520 | rs28416520 |
| rs7318091 | rs7318091 | rs7318091 | rs7318091 | rs7318091 |
| rs11571815 | rs11571815 | rs11571815 | — | — |
| rs112727324 | — | — | — | — |
| rs3736830 | rs3736830 | rs3736830 | rs3736830 | rs3736830 |
| rs7322160 | rs7322160 | rs7322160 | rs7322160 | rs7322160 |
| rs78566820 | rs78566820 | rs78566820 | — | — |
| rs12868295 | rs12868295 | rs12868295 | rs12868295 | rs12868295 |
| rs112097885 | — | — | — | — |
| rs1713426 | rs1713426 | rs1713426 | rs1713426 | rs1713426 |
| rs12879626 | rs12879626 | rs12879626 | rs12879626 | rs12879626 |
| rs61488898 | rs61488898 | rs61488898 | — | rs61488898 |
| rs45547534 | rs45547534 | rs45547534 | — | rs45547534 |
| rs1969713 | rs1969713 | rs1969713 | rs1969713 | rs1969713 |
| rs112326803 | rs112326803 | rs112326803 | — | rs112326803 |
| rs762643 | rs762643 | rs762643 | rs762643 | rs762643 |
| rs1986616 | rs1986616 | rs1986616 | rs1986616 | rs1986616 |
| rs71126035 | — | — | — | — |
| rs375453823 | — | — | — | — |
| rs9796 | rs9796 | — | — | — |
| rs689816 | rs689816 | rs689816 | rs689816 | rs689816 |
| rs11071756 | rs11071756 | rs11071756 | rs11071756 | rs11071756 |
| rs8025068 | rs8025068 | rs8025068 | rs8025068 | rs8025068 |
| rs8035724 | rs8035724 | rs8035724 | rs8035724 | rs8035724 |
| rs376442546 | — | — | — | — |
| rs716886 | rs716886 | rs716886 | rs716886 | rs716886 |
| rs62025270 | rs62025270 | rs62025270 | rs62025270 | rs62025270 |
| rs12898357 | rs12898357 | rs12898357 | rs12898357 | rs12898357 |
| rs28652789 | rs28652789 | rs28652789 | rs28652789 | rs28652789 |
| rs4786509 | rs4786509 | — | — | — |
| rs17680522 | rs17680522 | rs17680522 | rs17680522 | rs17680522 |
| rs8045589 | rs8045589 | — | — | — |
| rs9673473 | rs9673473 | — | — | — |
| rs2173885 | — | — | — | — |
| rs11642909 | rs11642909 | rs11642909 | rs11642909 | rs11642909 |
| rs11075466 | rs11075466 | rs11075466 | rs11075466 | rs11075466 |
| rs10521305 | rs10521305 | rs10521305 | — | rs10521305 |
| rs8045027 | rs8045027 | rs8045027 | rs8045027 | rs8045027 |
| rs200293726 | — | — | — | — |
| rs12447180 | rs12447180 | rs12447180 | rs12447180 | rs12447180 |
| rs502258 | rs502258 | rs502258 | rs502258 | rs502258 |
| rs62054610 | — | — | — | — |
| rs2108839 | — | — | — | — |
| rs5030755 | rs5030755 | rs5030755 | rs5030755 | rs5030755 |
| rs34856659 | rs34856659 | rs34856659 | rs34856659 | rs34856659 |
| rs222859 | — | — | — | — |
| rs569145577 | — | — | — | — |
| rs8073434 | rs8073434 | rs8073434 | rs8073434 | rs8073434 |
| rs1565920 | rs1565920 | rs1565920 | rs1565920 | rs1565920 |
| rs111637825 | rs111637825 | rs111637825 | — | rs111637825 |
| rs799903 | — | — | — | — |
| rs1815198 | rs1815198 | rs1815198 | rs1815198 | rs1815198 |
| rs1991401 | rs1991401 | rs1991401 | rs1991401 | rs1991401 |
| rs34609096 | rs34609096 | rs34609096 | rs34609096 | rs34609096 |
| rs4800141 | rs4800141 | rs4800141 | rs4800141 | rs4800141 |
| rs142239484 | rs142239484 | rs142239484 | — | — |
| rs12605881 | rs12605881 | — | — | — |
| rs11542373 | rs11542373 | rs11542373 | rs11542373 | rs11542373 |
| rs349306 | rs349306 | rs349306 | rs349306 | rs349306 |
| rs56225015 | rs56225015 | rs56225015 | rs56225015 | rs56225015 |
| rs12459191 | rs12459191 | rs12459191 | rs12459191 | rs12459191 |
| rs147068659 | — | — | — | — |
| rs148807049 | rs148807049 | rs148807049 | rs148807049 | rs148807049 |
| rs10406053 | rs10406053 | rs10406053 | rs10406053 | rs10406053 |
| rs191031532 | — | — | — | — |
| rs2359811 | rs2359811 | rs2359811 | rs2359811 | rs2359811 |
| rs11670032 | rs11670032 | rs11670032 | rs11670032 | rs11670032 |
| rs424223 | rs424223 | rs424223 | rs424223 | rs424223 |
| rs7249357 | rs7249357 | rs7249357 | rs7249357 | rs7249357 |
| rs11349039 | — | — | — | — |
| rs10409547 | — | — | — | — |
| rs11668344 | rs11668344 | rs11668344 | rs11668344 | rs11668344 |
| rs117146677 | rs117146677 | rs117146677 | — | rs117146677 |
| rs8109672 | rs8109672 | rs8109672 | rs8109672 | rs8109672 |
| rs12461110 | rs12461110 | rs12461110 | rs12461110 | rs12461110 |
| rs8101859 | rs8101859 | rs8101859 | rs8101859 | rs8101859 |
| rs394448 | rs394448 | — | — | — |
| rs6054257 | rs6054257 | rs6054257 | rs6054257 | rs6054257 |
| rs73078112 | rs73078112 | rs73078112 | — | rs73078112 |
| rs236117 | rs236117 | rs236117 | rs236117 | rs236117 |
| rs16991615 | rs16991615 | rs16991615 | — | rs16991615 |
| rs148563363 | rs148563363 | rs148563363 | — | — |
| rs746748 | rs746748 | rs746748 | — | rs746748 |
| rs58065489 | rs58065489 | rs58065489 | rs58065489 | rs58065489 |
| rs11699793 | rs11699793 | rs11699793 | rs11699793 | rs11699793 |
| rs483508 | rs483508 | rs483508 | rs483508 | rs483508 |
| rs80047570 | — | — | — | — |
| rs7266248 | rs7266248 | rs7266248 | rs7266248 | rs7266248 |
| rs73307923 | rs73307923 | rs73307923 | — | rs73307923 |
| rs6011452 | rs6011452 | rs6011452 | rs6011452 | rs6011452 |
| rs372721829 | rs372721829 | rs372721829 | — | — |
| rs10854167 | rs10854167 | rs10854167 | rs10854167 | rs10854167 |
| rs34978822 | rs34978822 | rs34978822 | — | rs34978822 |
| rs139337637 | — | — | — | — |
| rs9975728 | rs9975728 | rs9975728 | rs9975728 | rs9975728 |
| rs4148974 | rs4148974 | rs4148974 | — | rs4148974 |
| rs5746740 | rs5746740 | rs5746740 | rs5746740 | rs5746740 |
| rs5754100 | rs5754100 | rs5754100 | rs5754100 | rs5754100 |
| rs3838149 | — | — | — | — |
| rs62237617 | rs62237617 | rs62237617 | — | — |
| rs5762852 | rs5762852 | rs5762852 | rs5762852 | rs5762852 |
| rs140353819 | rs140353819 | rs140353819 | — | — |
| rs139906882 | rs139906882 | rs139906882 | — | rs139906882 |
| rs138430 | rs138430 | rs138430 | rs138430 | rs138430 |
| rs2958663 | rs2958663 | — | — | — |
| rs2272805 | rs2272805 | rs2272805 | rs2272805 | rs2272805 |
| rs148258173 | — | — | — | — |
| rs2382759 | — | — | — | — |
| rs62588628 | — | — | — | — |
| rs5953379 | — | — | — | — |
| rs5981360 | — | — | — | — |
| rs3213462 | — | — | — | — |

Abbreviations: GWAS, genome-wide association study; SNP, single nucleotide polymorphism. For more details on the SNPs see the GWAS by Ruth et al.

**eTable 2.** Association between age at menopause and risk of stroke in EPIC-CVD (n=7,883).

| Outcome/Age at menopause (years) | No. of cases | Model 1 |  | Model 2 |  | Model 3 |  |
| --- | --- | --- | --- | --- | --- | --- | --- |
|  |  | HR (95% CI) | P<sub>trend</sub> | HR (95% CI) | P<sub>trend</sub> | HR (95% CI) | P<sub>trend</sub> |
| Stroke |  |  |  |  |  |  |  |
| <40 | 240 | 1.44 (1.20-1.73) |  | 1.34 (1.10-1.63) |  | 1.37 (1.12-1.67) |  |
| 40 to<45 | 340 | 1.30 (1.11-1.53) |  | 1.24 (1.05-1.46) |  | 1.25 (1.06-1.47) |  |
| 45 to <50 | 828 | 1.16 (1.05-1.29) | <0.001 | 1.13 (1.02-1.26) | <0.001 | 1.14 (1.02-1.27) | <0.001 |
| 50 to <55 | 1,095 | 1.00 (0.92-1.08) |  | 1.00 (0.92-1.09) |  | 1.00 (0.92-1.09) |  |
| ≥55 | 235 | 0.88 (0.73-1.05) |  | 0.87 (0.72-1.05) |  | 0.88 (0.73-1.07) |  |
| per 5 years younger | 2,738 | 1.13 (1.08-1.18) |  | 1.10 (1.05-1.16) |  | 1.11 (1.06-1.16) |  |
| Ischemic stroke |  |  |  |  |  |  |  |
| <40 | 193 | 1.52 (1.24-1.85) |  | 1.40 (1.13-1.73) |  | 1.43 (1.15-1.77) |  |
| 40 to<45 | 270 | 1.34 (1.13-1.59) |  | 1.27 (1.06-1.52) |  | 1.28 (1.07-1.53) |  |
| 45 to <50 | 664 | 1.17 (1.04-1.30) | <0.001 | 1.14 (1.01-1.28) | <0.001 | 1.14 (1.02-1.28) | <0.001 |
| 50 to <55 | 874 | 1.00 (0.92-1.09) |  | 1.00 (0.91-1.10) |  | 1.00 (0.91-1.10) |  |
| ≥55 | 185 | 0.85 (0.70-1.04) |  | 0.84 (0.69-1.03) |  | 0.85 (0.70-1.05) |  |
| per 5 years younger | 2,186 | 1.14 (1.09-1.20) |  | 1.12 (1.06-1.17) |  | 1.12 (1.06-1.18) |  |
| Hemorrhagic stroke |  |  |  |  |  |  |  |
| <40 | 47 | 1.16 (0.84-1.60) |  | 1.10 (0.80-1.53) |  | 1.13 (0.81-1.57) |  |
| 40 to<45 | 70 | 1.15 (0.86-1.54) |  | 1.13 (0.84-1.52) |  | 1.13 (0.84-1.53) |  |
| 45 to <50 | 164 | 1.13 (0.95-1.35) | 0.232 | 1.11 (0.92-1.33) | 0.439 | 1.11 (0.93-1.33) | 0.384 |
| 50 to <55 | 221 | 1.00 (0.86-1.16) |  | 1.00 (0.85-1.17) |  | 1.00 (0.85-1.17) |  |
| ≥55 | 50 | 0.98 (0.71-1.35) |  | 1.02 (0.74-1.41) |  | 1.02 (0.74-1.41) |  |
| per 5 years younger | 552 | 1.07 (0.99-1.16) |  | 1.05 (0.97-1.14) |  | 1.06 (0.97-1.15) |  |
| Intracerebral hemorrhage |  |  |  |  |  |  |  |
| <40 | 30 | 1.31 (0.89-1.93) |  | 1.32 (0.89-1.96) |  | 1.34 (0.90-2.00) |  |
| 40 to<45 | 49 | 1.31 (0.92-1.88) |  | 1.32 (0.92-1.90) |  | 1.33 (0.93-1.91) |  |
| 45 to <50 | 91 | 1.01 (0.81-1.27) | 0.050 | 1.03 (0.81-1.30) | 0.054 | 1.03 (0.82-1.30) | 0.047 |
| 50 to <55 | 147 | 1.00 (0.83-1.20) |  | 1.00 (0.83-1.21) |  | 1.00 (0.83-1.21) |  |
| ≥55 | 28 | 0.79 (0.52-1.20) |  | 0.81 (0.53-1.22) |  | 0.80 (0.53-1.22) |  |
| per 5 years younger | 345 | 1.15 (1.04-1.26) |  | 1.14 (1.04-1.26) |  | 1.15 (1.04-1.27) |  |
| Subarachnoid hemorrhage |  |  |  |  |  |  |  |
| <40 | 17 | 0.93 (0.53-1.61) |  | 0.79 (0.44-1.41) |  | 0.81 (0.45-1.45) |  |
| 40 to<45 | 21 | 0.88 (0.54-1.44) |  | 0.83 (0.51-1.35) |  | 0.84 (0.51-1.36) |  |
| 45 to <50 | 73 | 1.32 (1.03-1.71) | 0.484 | 1.22 (0.94-1.58) | 0.182 | 1.22 (0.94-1.58) | 0.216 |
| 50 to <55 | 74 | 1.00 (0.77-1.29) |  | 1.00 (0.77-1.30) |  | 1.00 (0.77-1.30) |  |
| ≥55 | 22 | 1.44 (0.89-2.32) |  | 1.56 (0.95-2.55) |  | 1.55 (0.95-2.55) |  |
| per 5 years younger | 207 | 0.94 (0.82-1.09) |  | 0.89 (0.77-1.04) |  | 0.90 (0.78-1.05) |  |

Model 1 is adjusted for age; Model 2 is additionally adjusted for smoking status, body mass index, HbA1c, total cholesterol, and hypertension; Model 3 is additionally adjusted for ever use of hormone replacement therapy and age at menarche. Confidence intervals for each age at menopause category are presented using quasi variances in order to enhance the comparison between individual categories. Age at menopause from 50 to <55 years was used as reference category. P<sub>trend</sub> indicates P-value for linear trend. Abbreviations: CI, confidence interval; HR, hazard ratio.

**eTable 3.** Association between age at menopause and risk of stroke in the UK Biobank (n=196,361).

| Outcome/Age at menopause (years) | No. of cases | Model 1 |  | Model 2 |  | Model 3 |  |
| --- | --- | --- | --- | --- | --- | --- | --- |
|  |  | HR (95% CI) | P<sub>trend</sub> | HR (95% CI) | P<sub>trend</sub> | HR (95% CI) | P<sub>trend</sub> |
| Stroke |  |  |  |  |  |  |  |
| <40 | 460 | 1.63 (1.48-1.78) |  | 1.46 (1.33-1.61) |  | 1.45 (1.32-1.59) |  |
| 40 to<45 | 551 | 1.31 (1.20-1.42) |  | 1.23 (1.13-1.34) |  | 1.23 (1.13-1.33) |  |
| 45 to <50 | 914 | 1.11 (1.04-1.19) | <0.001 | 1.08 (1.01-1.16) | <0.001 | 1.08 (1.01-1.15) | <0.001 |
| 50 to <55 | 1,528 | 1.00 (0.95-1.05) |  | 1.00 (0.95-1.05) |  | 1.00 (0.95-1.06) |  |
| ≥55 | 579 | 1.02 (0.94-1.11) |  | 1.01 (0.93-1.10) |  | 1.01 (0.93-1.10) |  |
| per 5 years younger | 4,032 | 1.12 (1.09-1.15) |  | 1.09 (1.07-1.12) |  | 1.09 (1.06-1.12) |  |
| Ischemic stroke |  |  |  |  |  |  |  |
| <40 | 341 | 1.65 (1.48-1.84) |  | 1.45 (1.30-1.61) |  | 1.43 (1.28-1.59) |  |
| 40 to<45 | 383 | 1.25 (1.13-1.38) |  | 1.16 (1.05-1.29) |  | 1.15 (1.04-1.28) |  |
| 45 to <50 | 686 | 1.15 (1.06-1.24) | <0.001 | 1.11 (1.02-1.20) | <0.001 | 1.10 (1.02-1.19) | <0.001 |
| 50 to <55 | 1,121 | 1.00 (0.94-1.07) |  | 1.00 (0.94-1.07) |  | 1.00 (0.94-1.07) |  |
| ≥55 | 438 | 1.04 (0.94-1.15) |  | 1.02 (0.93-1.13) |  | 1.02 (0.93-1.12) |  |
| per 5 years younger | 2,969 | 1.12 (1.09-1.15) |  | 1.09 (1.06-1.12) |  | 1.08 (1.05-1.11) |  |
| Hemorrhagic stroke |  |  |  |  |  |  |  |
| <40 | 119 | 1.56 (1.30-1.87) |  | 1.52 (1.26-1.82) |  | 1.53 (1.27-1.84) |  |
| 40 to<45 | 168 | 1.47 (1.26-1.71) |  | 1.44 (1.24-1.68) |  | 1.45 (1.24-1.69) |  |
| 45 to <50 | 228 | 1.03 (0.90-1.17) | <0.001 | 1.01 (0.88-1.15) | <0.001 | 1.01 (0.89-1.16) | <0.001 |
| 50 to <55 | 407 | 1.00 (0.90-1.11) |  | 1.00 (0.90-1.11) |  | 1.00 (0.90-1.11) |  |
| ≥55 | 141 | 0.96 (0.81-1.14) |  | 0.97 (0.82-1.16) |  | 0.97 (0.82-1.16) |  |
| per 5 years younger | 1,063 | 1.13 (1.08-1.18) |  | 1.12 (1.07-1.17) |  | 1.12 (1.07-1.18) |  |
| Intracerebral hemorrhage |  |  |  |  |  |  |  |
| <40 | 72 | 1.62 (1.28-2.04) |  | 1.57 (1.25-1.99) |  | 1.60 (1.27-2.02) |  |
| 40 to<45 | 96 | 1.41 (1.15-1.73) |  | 1.39 (1.14-1.70) |  | 1.41 (1.15-1.72) |  |
| 45 to <50 | 133 | 1.02 (0.85-1.22) | <0.001 | 1.01 (0.85-1.20) | <0.001 | 1.01 (0.85-1.21) | <0.001 |
| 50 to <55 | 247 | 1.00 (0.88-1.14) |  | 1.00 (0.87-1.14) |  | 1.00 (0.87-1.15) |  |
| ≥55 | 83 | 0.86 (0.68-1.09) |  | 0.87 (0.69-1.10) |  | 0.87 (0.69-1.10) |  |
| per 5 years younger | 631 | 1.15 (1.08-1.22) |  | 1.14 (1.07-1.21) |  | 1.14 (1.07-1.21) |  |
| Subarachnoid hemorrhage |  |  |  |  |  |  |  |
| <40 | 47 | 1.48 (1.10-1.98) |  | 1.44 (1.07-1.93) |  | 1.43 (1.06-1.92) |  |
| 40 to<45 | 72 | 1.55 (1.22-1.97) |  | 1.52 (1.20-1.93) |  | 1.52 (1.19-1.93) |  |
| 45 to <50 | 95 | 1.04 (0.85-1.27) | 0.009 | 1.01 (0.83-1.24) | 0.023 | 1.01 (0.83-1.24) | 0.028 |
| 50 to <55 | 160 | 1.00 (0.85-1.17) |  | 1.00 (0.85-1.17) |  | 1.00 (0.85-1.18) |  |
| ≥55 | 58 | 1.12 (0.86-1.45) |  | 1.15 (0.89-1.50) |  | 1.15 (0.88-1.50) |  |
| per 5 years younger | 432 | 1.10 (1.02-1.19) |  | 1.09 (1.01-1.18) |  | 1.09 (1.00-1.17) |  |

Model 1 is adjusted for age; Model 2 is additionally adjusted for smoking status, body mass index, HbA1c, total cholesterol, and hypertension; Model 3 is additionally adjusted for ever use of hormone replacement therapy and age at menarche. Confidence intervals for each age at menopause category are presented using quasi variances in order to enhance the comparison between individual categories. Age at menopause from 50 to <55 years was used as reference category. P<sub>trend</sub> indicates P-value for linear trend. Abbreviations: CI, confidence interval; HR, hazard ratio.

**eTable 4.** Mendelian Randomization analysis on genetically proxied age at menopause and risk of stroke.

| Method/Outcome | Overall |  | EPIC-CVD |  | UK Biobank |  |
| --- | --- | --- | --- | --- | --- | --- |
|  | HR (95% CI) | P-value | HR (95% CI) | P-value | HR (95% CI) | P-value |
| **Stroke** |  |  |  |  |  |  |
| IVW | 0.95 (0.82-1.09) | 0.445 | 1.01 (0.72-1.41) | 0.961 | 0.93 (0.80-1.09) | 0.386 |
| Simple median | 1.00 (0.79-1.25) | 0.982 | 1.00 (0.58-1.71) | 0.993 | 1.00 (0.77-1.29) | 0.983 |
| Weighted median | 0.98 (0.78-1.25) | 0.893 | 1.11 (0.63-1.94) | 0.717 | 0.96 (0.74-1.25) | 0.750 |
| MR-Egger | 0.91 (0.66-1.27) | 0.589 | 0.82 (0.38-1.77) | 0.611 | 0.94 (0.65-1.34) | 0.720 |
| Intercept | 1.00 (0.99-1.01) | 0.816 | 1.01 (0.99-1.03) | 0.556 | 1.00 (0.99-1.01) | 0.984 |
| **Ischemic stroke** |  |  |  |  |  |  |
| IVW | 0.98 (0.83-1.16) | 0.805 | 1.08 (0.75-1.57) | 0.672 | 0.96 (0.79-1.15) | 0.627 |
| Simple median | 0.99 (0.77-1.28) | 0.952 | 1.10 (0.62-1.97) | 0.738 | 0.97 (0.72-1.29) | 0.816 |
| Weighted median | 1.06 (0.81-1.39) | 0.681 | 1.25 (0.68-2.29) | 0.475 | 1.02 (0.75-1.38) | 0.920 |
| MR-Egger | 1.01 (0.69-1.48) | 0.960 | 0.87 (0.37-2.04) | 0.744 | 1.05 (0.68-1.61) | 0.827 |
| Intercept | 1.00 (0.99-1.01) | 0.862 | 1.01 (0.98-1.03) | 0.570 | 1.00 (0.99-1.01) | 0.633 |
| **Hemorrhagic stroke** |  |  |  |  |  |  |
| IVW | 0.85 (0.65-1.11) | 0.230 | 0.77 (0.44-1.37) | 0.377 | 0.87 (0.64-1.18) | 0.374 |
| Simple median | 0.98 (0.64-1.52) | 0.940 | 1.04 (0.42-2.58) | 0.935 | 0.97 (0.59-1.59) | 0.896 |
| Weighted median | 0.86 (0.55-1.34) | 0.508 | 0.81 (0.31-2.15) | 0.679 | 0.87 (0.53-1.44) | 0.594 |
| MR-Egger | 0.68 (0.37-1.26) | 0.219 | 0.70 (0.19-2.58) | 0.593 | 0.67 (0.33-1.36) | 0.268 |
| Intercept | 1.01 (0.99-1.02) | 0.432 | 1.00 (0.97-1.04) | 0.867 | 1.01 (0.99-1.03) | 0.422 |
| **Intracerebral hemorrhage** |  |  |  |  |  |  |
| IVW | 0.92 (0.65-1.29) | 0.617 | 0.90 (0.44-1.83) | 0.770 | 0.92 (0.62-1.37) | 0.683 |
| Simple median | 1.00 (0.57-1.76) | 0.990 | 1.20 (0.37-3.89) | 0.756 | 0.95 (0.50-1.80) | 0.877 |
| Weighted median | 1.11 (0.62-2.00) | 0.715 | 1.28 (0.38-4.26) | 0.691 | 1.07 (0.55-2.08) | 0.843 |
| MR-Egger | 0.97 (0.44-2.15) | 0.935 | 0.74 (0.14-3.78) | 0.713 | 1.05 (0.42-2.62) | 0.911 |
| Intercept | 1.00 (0.98-1.02) | 0.880 | 1.01 (0.97-1.05) | 0.790 | 1.00 (0.97-1.02) | 0.748 |
| **Subarachnoid hemorrhage** |  |  |  |  |  |  |
| IVW | 0.75 (0.50-1.14) | 0.184 | 0.61 (0.26-1.43) | 0.252 | 0.81 (0.50-1.30) | 0.377 |
| Simple median | 0.80 (0.41-1.55) | 0.503 | 0.56 (0.15-2.08) | 0.383 | 0.90 (0.42-1.95) | 0.790 |
| Weighted median | 0.64 (0.33-1.24) | 0.187 | 0.56 (0.14-2.19) | 0.403 | 0.67 (0.31-1.43) | 0.296 |
| MR-Egger | 0.40 (0.15-1.06) | 0.066 | 0.62 (0.09-4.49) | 0.640 | 0.35 (0.12-1.06) | 0.064 |
| Intercept | 1.02 (0.99-1.04) | 0.161 | 1.00 (0.95-1.05) | 0.974 | 1.02 (1.00-1.05) | 0.104 |

Abbreviations: CI, confidence interval; IVW, inverse variance weighted; HR, hazard ratio. HRs are per five years younger genetically proxied age at menopause.

**eTable 5.** Stroke risk factors and female-specific factors stratified by fifths of the polygenic risk score for age at menopause.

| Characteristic | Polygenic risk score* |  |  |  |  |
| --- | --- | --- | --- | --- | --- |
|  | >0.393 to 1.12<br>n=39,128 | >0.275 to 0.393<br>n=39,127 | >0.172 to 0.275<br>n=39,127 | >0.0537 to 0.172<br>n=39,127 | -0.675 to 0.0537<br>n=39,127 |
| **Stroke risk factors** |  |  |  |  |  |
| Age, years | 58.8 ± 6.0 | 59.4 ± 5.8 | 59.3 ± 5.7 | 59.6 ± 5.6 | 59.8 ± 5.5 |
| Hypertension | 19,466 (49.8%) | 19,946 (51.0%) | 20,012 (51.2%) | 20,401 (52.2%) | 20,771 (53.1%) |
| Body mass index, kg/m<sup>2</sup> | 26.8 ± 5.1 | 26.8 ± 5.0 | 26.9 ± 5.0 | 26.6 ± 5.1 | 26.8 ± 5.1 |
| Smoking status |  |  |  |  |  |
| Never | 22,352 (57.4%) | 22,544 (57.9%) | 22,501 (57.8%) | 22,596 (58.1%) | 22,687 (58.3%) |
| Ex | 13,169 (33.8%) | 12,935 (33.2%) | 12,990 (33.4%) | 12,868 (33.1%) | 12,865 (33.1%) |
| Current | 3,419 (8.8%) | 3,451 (8.9%) | 3,435 (8.8%) | 3,457 (8.9%) | 3,371 (8.7%) |
| Total cholesterol, mmol/L | 6.2 ± 1.1 | 6.2 ± 1.1 | 6.2 ± 1.1 | 6.2 ± 1.1 | 6.2 ± 1.1 |
| HbA1c, % | 5.6 ± 0.6 | 5.5 ± 0.5 | 5.5 ± 0.5 | 5.5 ± 0.6 | 5.6 ± 0.6 |
| **Female-specific factors** |  |  |  |  |  |
| Ever use of HRT | 20,485 (52.7%) | 19,770 (50.8%) | 19,355 (49.8%) | 19,102 (49.1%) | 18,527 (47.7%) |
| Age at menarche, years | 13.1 ± 1.6 | 13.2 ± 1.6 | 13.2 ± 1.6 | 13.2 ± 1.6 | 13.3 ± 1.6 |

\*A higher polygenic risk score indicates a lower genetically proxied age at menopause. The denominator for all percentages was the number of postmenopausal women without missing values in the respective variable. Abbreviations: HbA1c, glycated hemoglobin; HRT, hormone replacement therapy.

**eTable 6.** Stroke risk factors and female-specific factors stratified by categories of age at menopause.

| Characteristic | Age at menopause, years |  |  |  |  |
| --- | --- | --- | --- | --- | --- |
|  | <40<br>n=17,461 | 40 to <45<br>n=23,508 | 45 to <50<br>n=45,778 | 50 to <55<br>n=77,933 | ≥55<br>n=24,415 |
| **Stroke risk factors** |  |  |  |  |  |
| Age, years | 55.4 ± 7.4 | 57.5 ± 6.9 | 58.4 ± 6.1 | 60.1 ± 4.9 | 62.3 ± 3.6 |
| Hypertension | 8,848 (50.9%) | 11,862 (50.7%) | 22,156 (48.6%) | 39,291 (50.6%) | 14,111 (57.9%) |
| Body mass index, kg/m<sup>2</sup> | 27.5 ± 5.5 | 27.0 ± 5.2 | 26.8 ± 5.1 | 26.7 ± 4.9 | 27.2 ± 5.0 |
| Smoking status |  |  |  |  |  |
| Never | 9,238 (53.1%) | 12,848 (54.9%) | 25,867 (56.7%) | 46,662 (60.1%) | 14,657 (60.3%) |
| Ex | 5,686 (32.7%) | 7,943 (33.9%) | 15,137 (33.2%) | 25,549 (32.9%) | 8,388 (34.5%) |
| Current | 2,463 (14.2%) | 2,612 (11.2%) | 4,590 (10.1%) | 5,489 (7.1%) | 1,270 (5.2%) |
| Total cholesterol, mmol/L | 6.0 ± 1.2 | 6.2 ± 1.2 | 6.2 ± 1.1 | 6.2 ± 1.1 | 6.3 ± 1.1 |
| HbA1c, % | 5.5 ± 0.7 | 5.5 ± 0.6 | 5.6 ± 0.5 | 5.6 ± 0.5 | 5.6 ± 0.5 |
| **Female-specific factors** |  |  |  |  |  |
| Ever use of HRT | 11,733 (67.6%) | 14,204 (60.8%) | 22,551 (49.5%) | 32,117 (41.4%) | 11,898 (48.9%) |
| Age at menarche, years | 12.9 ± 1.8 | 13.0 ± 1.7 | 13.1 ± 1.6 | 13.2 ± 1.6 | 13.4 ± 1.6 |

The denominator for all percentages was the number of postmenopausal women without missing values in the respective variable. Abbreviations: HbA1c, glycated hemoglobin; HRT, hormone replacement therapy.

### Supplemental Figures

**eFigure 1.**
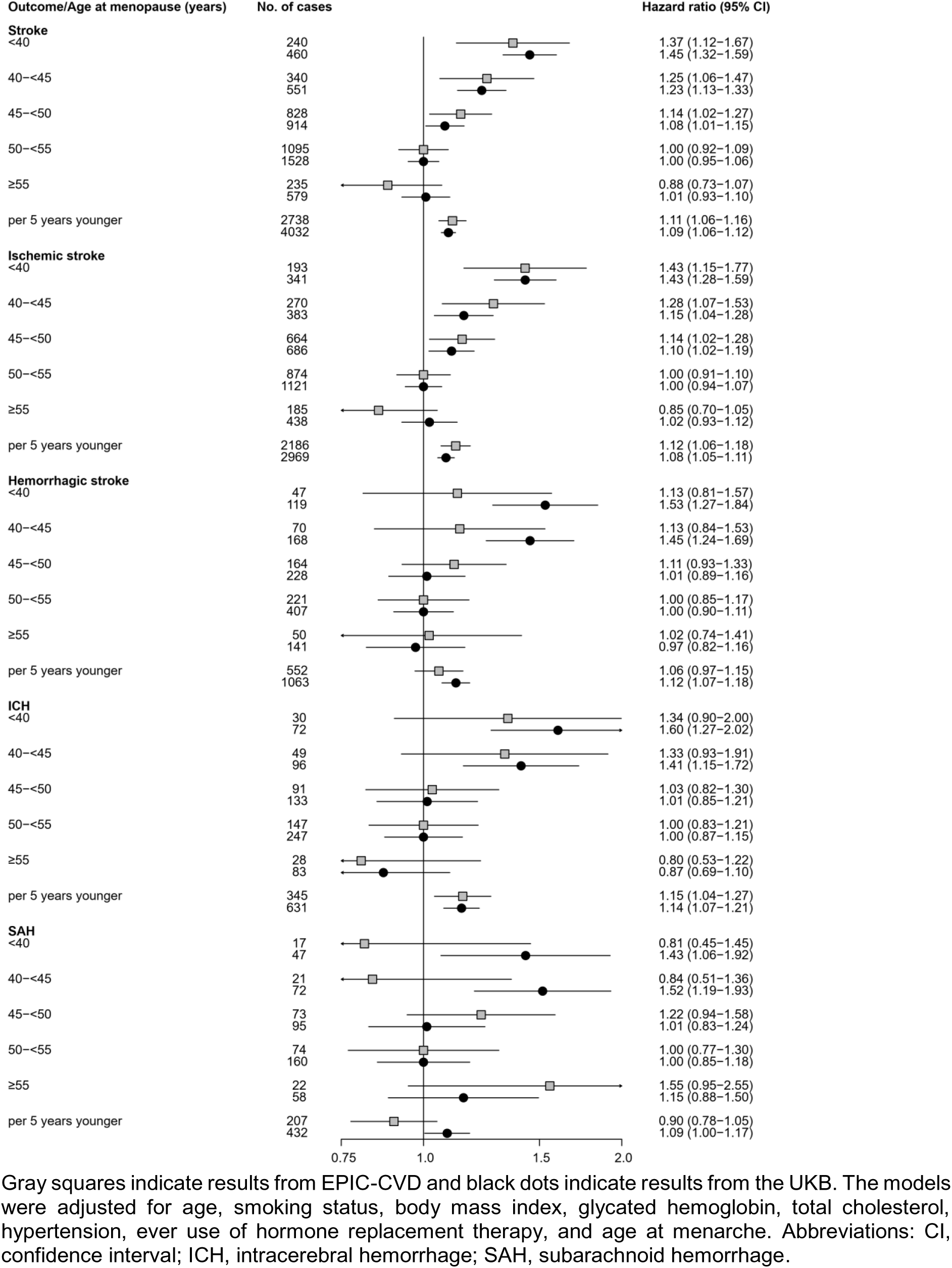
Consistency of associations between age at menopause and risk of various subtypes of stroke in EPIC-CVD and the UK Biobank.

**eFigure 2.**
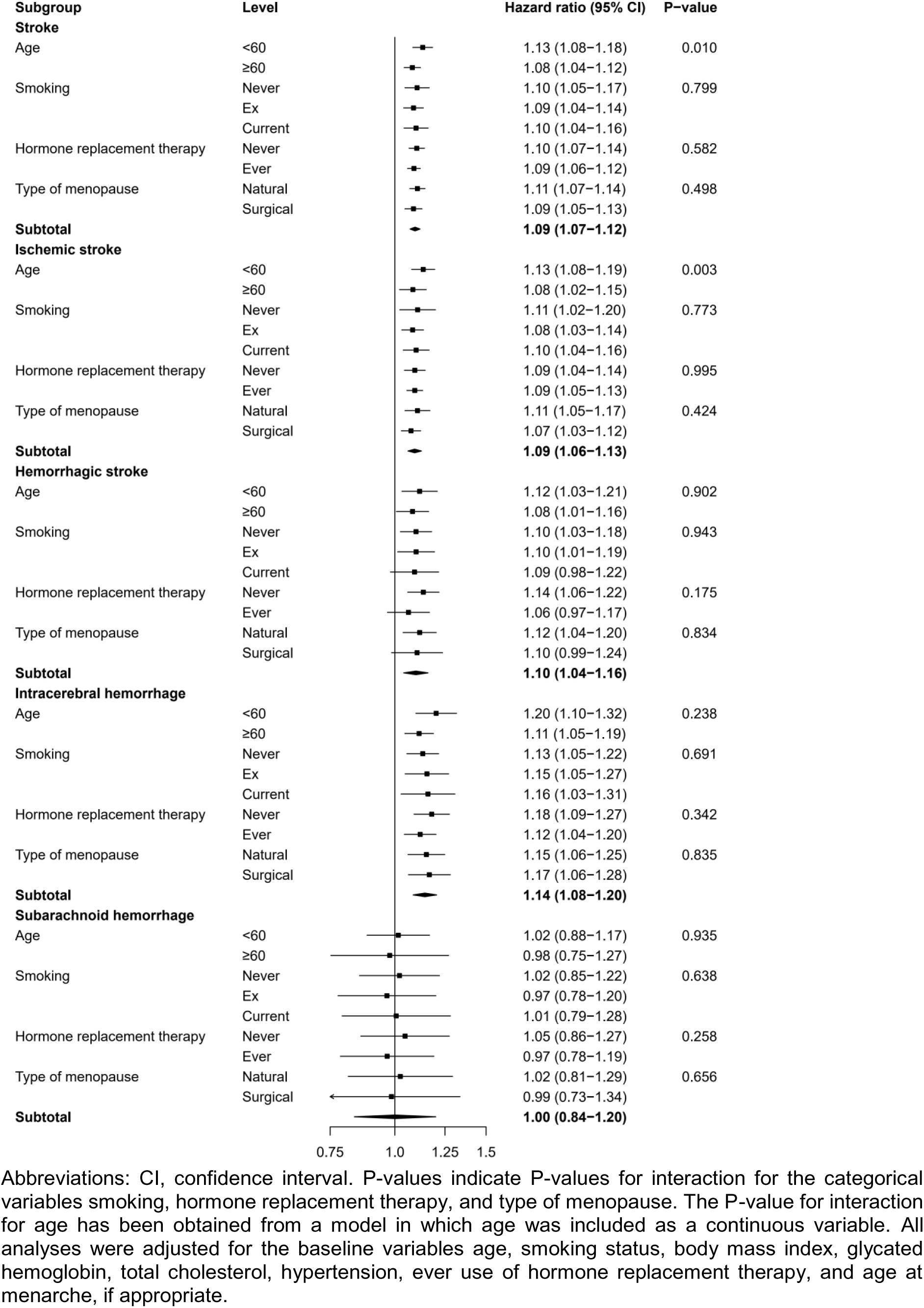
Observational analyses investigating the risk of different types of stroke per five years younger age at menopause across several subgroups.

**eFigure 3.**
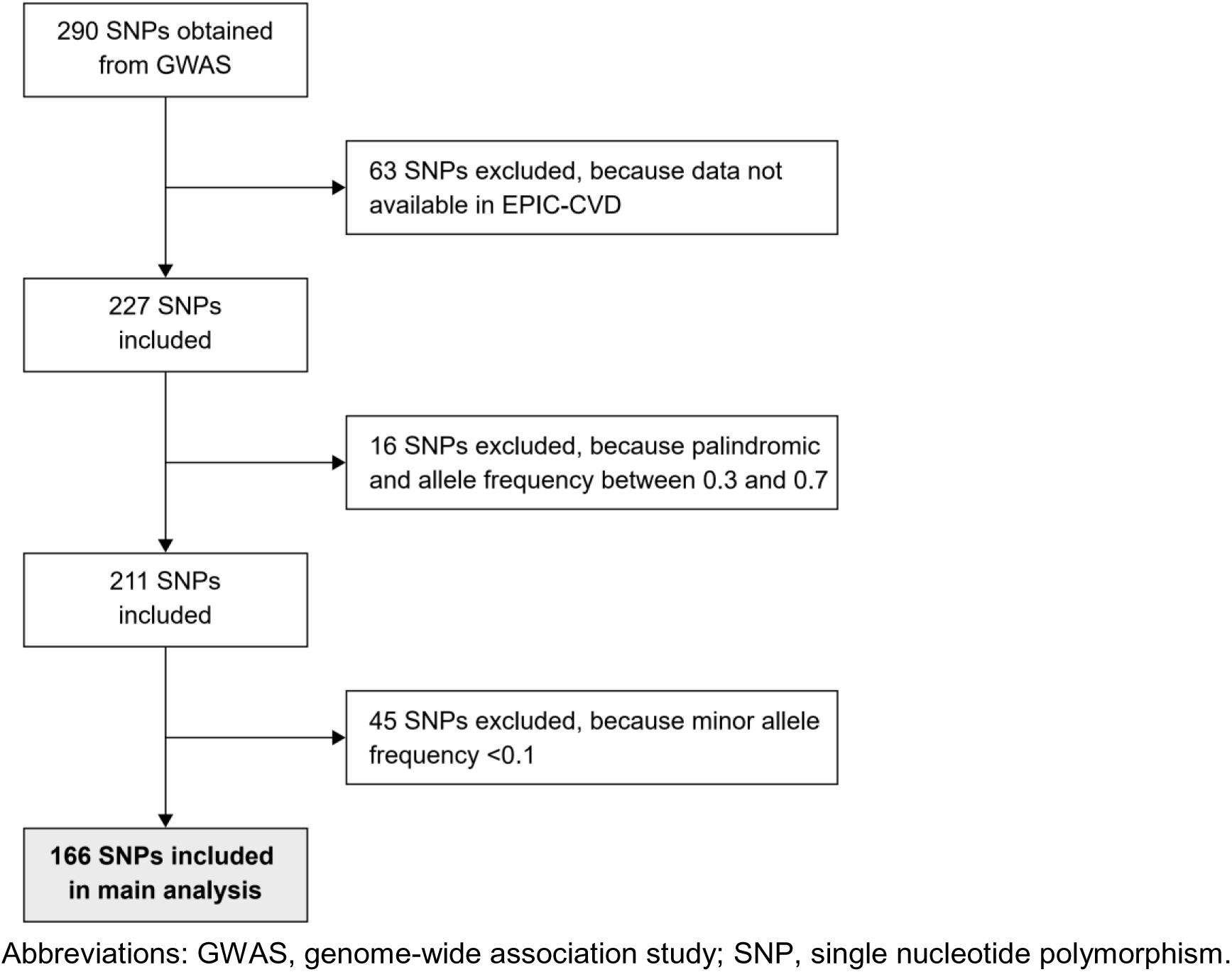
Flow chart for selection process of SNPs.

**eFigure 4.**
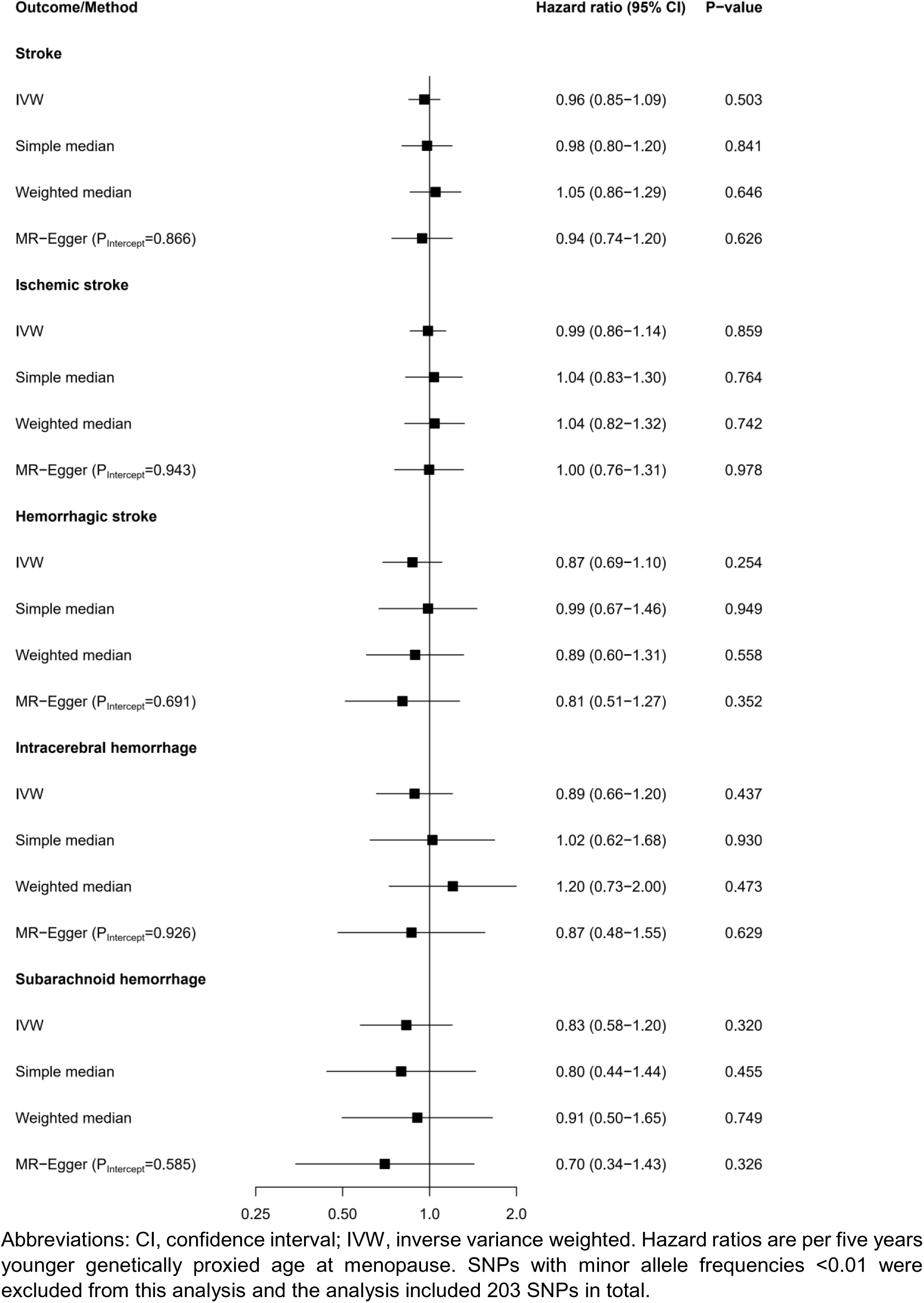
Mendelian Randomization analysis on genetically proxied age at menopause and risk of stroke excluding very rare genetic variants only.

**eFigure 5.**
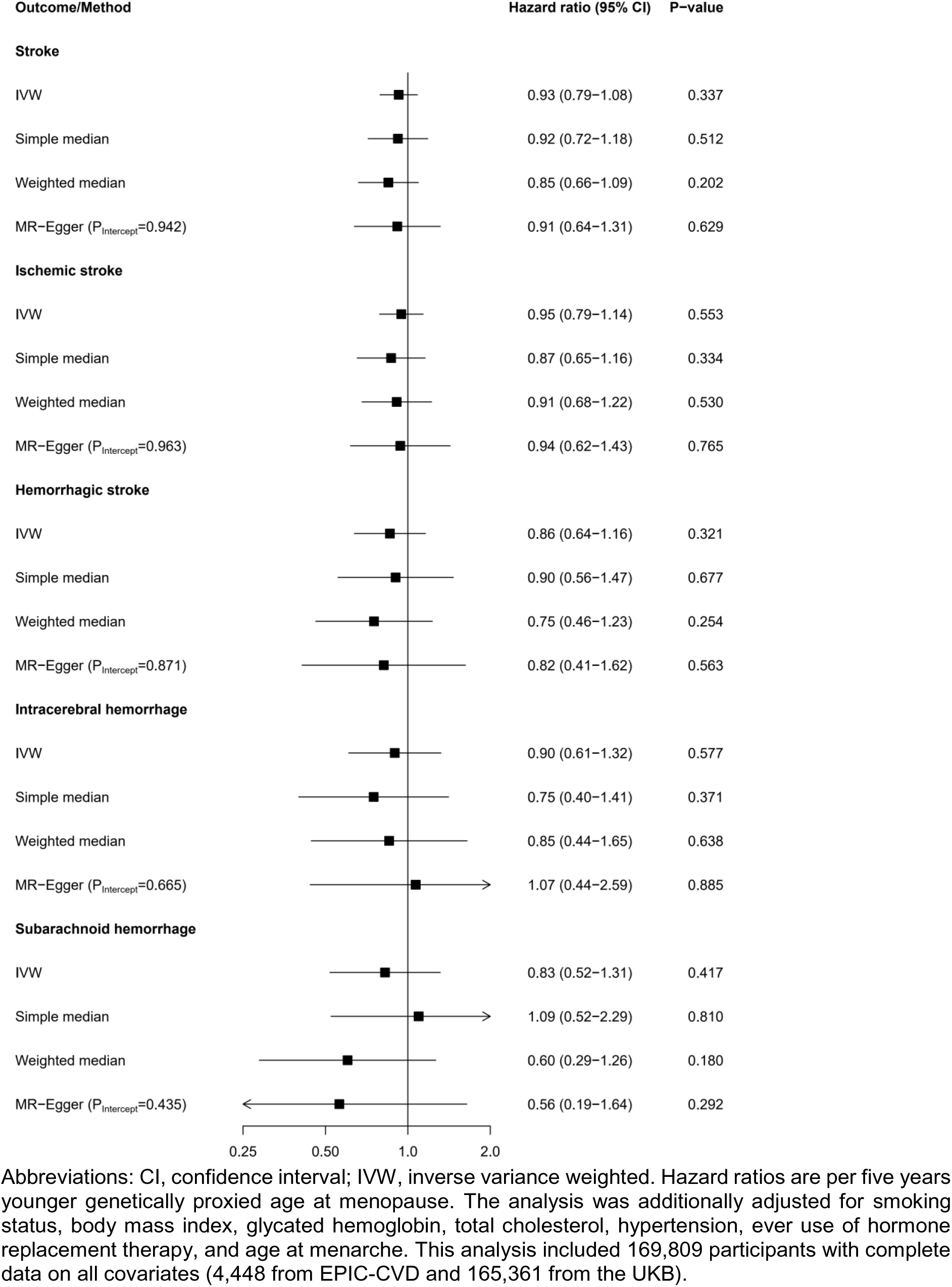
Mendelian Randomization analysis on genetically proxied age at menopause and risk of stroke adjusted for phenotypes related to cardiovascular risk.

## References

1. Vos T, Lim SS, Abbafati C, Abbas KM, Abbasi M, Abbasifard M, Abbasi-Kangevari M, Abbastabar H, Abd-Allah F, Abdelalim A, et al. Global burden of 369 diseases and injuries in 204 countries and territories, 1990–2019: a systematic analysis for the Global Burden of Disease Study 2019. Lancet. 2020;396:1204–1222. doi: 10.1016/S0140-6736(20)30925-9.

2. Institute for Health Metrics and Evaluation. GBD Compare Data Visualization. Seattle, WA: IHME, University of Washington web site. https://vizhub.healthdata.org/gbd-compare/. Accessed July 21, 2022.

3. Vyas MV, Silver FL, Austin PC, Yu AYX, Pequeno P, Fang J, Laupacis A, Kapral MK. Stroke Incidence by Sex Across the Lifespan. Stroke. 2021;52:447–451. doi: 10.1161/strokeaha.120.032898.

4. Rexrode KM, Madsen TE, Yu AYX, Carcel C, Lichtman JH, Miller EC. The Impact of Sex and Gender on Stroke. Circ Res. 2022;130:512–528. doi: 10.1161/CIRCRESAHA.121.319915.

5. O’Kelly AC, Michos ED, Shufelt CL, Vermunt JV, Minissian MB, Quesada O, Smith GN, Rich-Edwards JW, Garovic VD, El Khoudary SR, et al. Pregnancy and Reproductive Risk Factors for Cardiovascular Disease in Women. Circ Res. 2022;130:652–672. doi: 10.1161/CIRCRESAHA.121.319895.

6. Nelson HD. Menopause. Lancet. 2008;371:760–770. doi: 10.1016/S0140-6736(08)60346-3.

7. El Khoudary SR, Aggarwal B, Beckie TM, Hodis HN, Johnson AE, Langer RD, Limacher MC, Manson JE, Stefanick ML, Allison MA. Menopause Transition and Cardiovascular Disease Risk: Implications for Timing of Early Prevention: A Scientific Statement From the American Heart Association. Circulation. 2020;142:e506–e532. doi: 10.1161/CIR.0000000000000912.

8. Peters SA, Woodward M. Women’s reproductive factors and incident cardiovascular disease in the UK Biobank. Heart. 2018;104:1069–1075. doi: 10.1136/heartjnl-2017-312289.

9. Zhu D, Chung H-F, Dobson AJ, Pandeya N, Giles GG, Bruinsma F, Brunner EJ, Kuh D, Hardy R, Avis NE, et al. Age at natural menopause and risk of incident cardiovascular disease: a pooled analysis of individual patient data. The Lancet Public Health. 2019;4:e553–e564. doi: 10.1016/S2468-2667(19)30155-0.

10. Dam V, Onland-Moret NC, Burgess S, Chirlaque M-D, Peters SAE, Schuit E, Tikk K, Weiderpass E, Oliver-Williams C, Wood AM, et al. Genetically Determined Reproductive Aging and Coronary Heart Disease: A Bidirectional 2-sample Mendelian Randomization. J Clin Endocrinol Metab. 2022;107:e2952–e2961. doi: 10.1210/clinem/dgac171.

11. Ruth KS, Day FR, Hussain J, Martínez-Marchal A, Aiken CE, Azad A, Thompson DJ, Knoblochova L, Abe H, Tarry-Adkins JL, et al. Genetic insights into biological mechanisms governing human ovarian ageing. Nature. 2021;596:393–397. doi: 10.1038/s41586-021-03779-7.

12. Lankester J, Li J, Salfati ELI, Stefanick ML, Chan KHK, Liu S, Crandall CJ, Clarke SL, Assimes TL. Genetic evidence for causal relationships between age at natural menopause and the risk of ageing-associated adverse health outcomes. Int J Epidemiol. 2022. doi: 10.1093/ije/dyac215.

13. Allen N, Sudlow C, Downey P, Peakman T, Danesh J, Elliott P, Gallacher J, Green J, Matthews P, Pell J, et al. UK Biobank: Current status and what it means for epidemiology. Health Policy and Technology. 2012;1:123–126. doi: 10.1016/j.hlpt.2012.07.003.

14. Sudlow C, Gallacher J, Allen N, Beral V, Burton P, Danesh J, Downey P, Elliott P, Green J, Landray M, et al. UK biobank: an open access resource for identifying the causes of a wide range of complex diseases of middle and old age. PLoS Med. 2015;12:e1001779. doi: 10.1371/journal.pmed.1001779.

15. Riboli E, Hunt KJ, Slimani N, Ferrari P, Norat T, Fahey M, Charrondière UR, Hémon B, Casagrande C, Vignat J, et al. European Prospective Investigation into Cancer and Nutrition (EPIC): study populations and data collection. Public Health Nutr. 2002;5:1113–1124. doi: 10.1079/PHN2002394.

16. Danesh J, Saracci R, Berglund G, Feskens E, Overvad K, Panico S, Thompson S, Fournier A, Clavel-Chapelon F, Canonico M, et al. EPIC-Heart: the cardiovascular component of a prospective study of nutritional, lifestyle and biological factors in 520,000 middle-aged participants from 10 European countries. Eur J Epidemiol. 2007;22:129–141. doi: 10.1007/s10654-006-9096-8.

17. Langenberg C, Sharp S, Forouhi NG, Franks PW, Schulze MB, Kerrison N, Ekelund U, Barroso I, Panico S, Tormo MJ, et al. Design and cohort description of the InterAct Project: an examination of the interaction of genetic and lifestyle factors on the incidence of type 2 diabetes in the EPIC Study. Diabetologia. 2011;54:2272–2282. doi: 10.1007/s00125-011-2182-9.

18. Zheng J-S, Luan J, Sofianopoulou E, Imamura F, Stewart ID, Day FR, Pietzner M, Wheeler E, Lotta LA, Gundersen TE, et al. Plasma Vitamin C and Type 2 Diabetes: Genome-Wide Association Study and Mendelian Randomization Analysis in European Populations. Diabetes Care. 2021;44:98–106. doi: 10.2337/dc20-1328.

19. Collins R. What makes UK Biobank special? Lancet. 2012;379:1173–1174. doi: 10.1016/S0140-6736(12)60404-8.

20. Bycroft C, Freeman C, Petkova D, Band G, Elliott LT, Sharp K, Motyer A, Vukcevic D, Delaneau O, O’Connell J, et al. Genome-wide genetic data on ∼500,000 UK Biobank participants. bioRxiv. 2017. doi: 10.1101/166298.

21. Onland-Moret NC, van der A DL, van der Schouw YT, Buschers W, Elias SG, van Gils CH, Koerselman J, Roest M, Grobbee DE, Peeters PHM. Analysis of case-cohort data: a comparison of different methods. J Clin Epidemiol. 2007;60:350–355. doi: 10.1016/j.jclinepi.2006.06.022.

22. Firth D. Quasi-variances. Biometrika. 2004;91:65–80. doi: 10.1093/biomet/91.1.65.

23. White IR. Multivariate Random-effects Meta-analysis. Stata J. 2009;9:40–56. doi: 10.1177/1536867X0900900103.

24. Privé F, Luu K, Blum MGB, McGrath JJ, Vilhjálmsson BJ. Efficient toolkit implementing best practices for principal component analysis of population genetic data. Bioinformatics. 2020;36:4449–4457. doi: 10.1093/bioinformatics/btaa520.

25. Yavorska OO, Burgess S. MendelianRandomization: an R package for performing Mendelian randomization analyses using summarized data. Int J Epidemiol. 2017;46:1734–1739. doi: 10.1093/ije/dyx034.

26. Verbanck M, Chen C-Y, Neale B, Do R. Detection of widespread horizontal pleiotropy in causal relationships inferred from Mendelian randomization between complex traits and diseases. Nat Genet. 2018;50:693–698. doi: 10.1038/s41588-018-0099-7.

27. Dam V, van der Schouw YT, Onland-Moret NC, Groenwold RHH, Peters SAE, Burgess S, Wood AM, Chirlaque M-D, Moons KGM, Oliver-Williams C, et al. Association of menopausal characteristics and risk of coronary heart disease: a pan-European case-cohort analysis. Int J Epidemiol. 2019;48:1275–1285. doi: 10.1093/ije/dyz016.

28. Woodfield R, Grant I, Sudlow CLM. Accuracy of Electronic Health Record Data for Identifying Stroke Cases in Large-Scale Epidemiological Studies: A Systematic Review from the UK Biobank Stroke Outcomes Group. PLoS One. 2015;10:e0140533. doi: 10.1371/journal.pone.0140533.

29. Staiger D, Stock JH. Instrumental Variables Regression with Weak Instruments. Econometrica. 1997;65:557. doi: 10.2307/2171753.

30. Jung KJ, Kim M-R, Yun YD, Kim HC, Jee SH. Duration of ovarian hormone exposure and atherosclerotic cardiovascular disease in Korean women: the Korean Heart Study. Menopause. 2016;23:60–66. doi: 10.1097/GME.0000000000000489.

31. Yang L, Lin L, Kartsonaki C, Guo Y, Chen Y, Bian Z, Xie K, Jin D, Li L, Lv J, et al. Menopause Characteristics, Total Reproductive Years, and Risk of Cardiovascular Disease Among Chinese Women. Circ Cardiovasc Qual Outcomes. 2017;10. doi: 10.1161/CIRCOUTCOMES.117.004235.

32. Ley SH, Li Y, Tobias DK, Manson JE, Rosner B, Hu FB, Rexrode KM. Duration of Reproductive Life Span, Age at Menarche, and Age at Menopause Are Associated With Risk of Cardiovascular Disease in Women. J Am Heart Assoc. 2017;6. doi: 10.1161/JAHA.117.006713.

33. Lisabeth LD, Beiser AS, Brown DL, Murabito JM, Kelly-Hayes M, Wolf PA. Age at natural menopause and risk of ischemic stroke: the Framingham heart study. Stroke. 2009;40:1044–1049. doi: 10.1161/STROKEAHA.108.542993.

34. Gallagher LG, Davis LB, Ray RM, Psaty BM, Gao DL, Checkoway H, Thomas DB. Reproductive history and mortality from cardiovascular disease among women textile workers in Shanghai, China. Int J Epidemiol. 2011;40:1510–1518. doi: 10.1093/ije/dyr134.

35. Lai PMR, Jimenez M, Du R, Rexrode K. Association of Reproductive Life Span and Age at Menopause With the Risk of Aneurysmal Subarachnoid Hemorrhage. Neurology. 2022;98:e2005–e2012. doi: 10.1212/WNL.0000000000200222.

36. Iorga A, Cunningham CM, Moazeni S, Ruffenach G, Umar S, Eghbali M. The protective role of estrogen and estrogen receptors in cardiovascular disease and the controversial use of estrogen therapy. Biol Sex Differ. 2017;8:33. doi: 10.1186/s13293-017-0152-8.

37. Peters SAE, Woodward M. Oestradiol and the risk of myocardial infarction in women: a cohort study of UK Biobank participants. Int J Epidemiol. 2021;50:1241–1249. doi: 10.1093/ije/dyaa284.

38. Geistanger A, Arends S, Berding C, Hoshino T, Jeppsson J-O, Little R, Siebelder C, Weykamp C. Statistical methods for monitoring the relationship between the IFCC reference measurement procedure for hemoglobin A1c and the designated comparison methods in the United States, Japan, and Sweden. Clin Chem. 2008;54:1379–1385. doi: 10.1373/clinchem.2008.103556.

39. Peters T, Brage S, Westgate K, Franks PW, Gradmark A, Tormo Diaz MJ, Huerta JM, Bendinelli B, Vigl M, Boeing H, et al. Validity of a short questionnaire to assess physical activity in 10 European countries. Eur J Epidemiol. 2012;27:15–25. doi: 10.1007/s10654-011-9625-y.

40. Guidelines for data processing analysis of the International Physical Activity Questionnaire (IPAQ) - Short and long forms.

41. White IR, Royston P. Imputing missing covariate values for the Cox model. Stat Med. 2009;28:1982–1998. doi: 10.1002/sim.3618.

42. Burgess S, White IR, Resche-Rigon M, Wood AM. Combining multiple imputation and meta-analysis with individual participant data. Stat Med. 2013;32:4499– 4514. doi: 10.1002/sim.5844.

